# Using single-subject morphological networks to elucidate the patterns of disconnection and disconnectome associated with post-stroke deficits and recovery

**DOI:** 10.1101/2024.12.31.24319803

**Authors:** Binke Yuan, Tao Zhong, Yaling Wang, Qingwen Chen, Xiaolin Guo, Junjie Yang, Xiaowei Gao, Zhe Hu, Junjing Li, Jiaxuan Liu, Zhiheng Qu, Wanchun Li, Zhongqi Li, Wanjing Li, Yien Huang, Jiali Chen, Hao Wen, Ye Zhang, Junle Li, Han Gao

## Abstract

**BACKGROUND:** The single-subject morphological network (SSMN) provides a new approach for constructing structural connectome. However, its clinical relevance in post-stroke deficits and recovery remains unexplored.

**METHODS:** This study utilized high-resolution 3D T1-weighted images alongside behavioral and cognitive assessments across multiple domains, including language, motor, memory, and attention, collected at two weeks, three months, and one year post-stroke. The SSMN was constructed using the AAL atlas by evaluating the similarities of regional probability density derived from gray matter volume. Network disconnection and the disconnectome were evaluated by examining changes in network edges and global topological properties. The functional relevance of the SSMN was explored through its associations with post-stroke behavioral and cognitive deficits and recovery, as well as by developing machine-learning-based prediction models.

**RESULTS:** The findings revealed that the SSMN was sensitive to post-stroke connectional and connectomal disruptions. Domain-specific disruptions in the SSMN were predictable of post-stroke deficits, with correlation pattern aligning with the neurobiological substrates of each domain. Furthermore, the predictive performance of SSMN-based models was comparable to that of other imaging modalities. Notably, normalization of the SSMN within one year post-stroke was significantly associated with functional recovery.

**CONCLUSIONS:** These results highlight the potential of the SSMN as a novel structural imaging modality for evaluating post-stroke deficits and recovery, offering valuable insights into the neurobiological mechanisms of rehabilitation.

## Introduction

Stroke is one of the leading causes of death and long-term disability worldwide [1]. Stroke resulted in a range of behavioral and cognitive deficits [2] by focal brain cell death and disruption of axonal pathways [3, 4]. In connectome neuroscience, the brain is conceptualized as a structural or functional wiring diagram that connects various regions [5, 6]. Numerous neuroimaging studies have found that stroke induced widespread disruptions in connectivity and network topology [7, 8], collectively referred to as the disconnectome [9, 10]. To investigate the disconnectome after stroke, previous research has predominantly relied on diffusion-weighted imaging tractography and resting-state fMRI [8, 11–16]. While structural MRI using 3D T1-weighted imaging has traditionally been employed to analyze local morphological features, such as gray matter volume (GMV) and cortical thickness [17, 18], recent methodological advancements have enabled the construction of the human connectome using single-subject 3D T1 images [19–22]. These developments open new avenues for exploring stroke-induced network disruptions.

Single-Subject Morphological Network (SSMN) refers to construct brain connectome by calculating the pairwise statistical interdependence of local morphological features from single-subject 3D T1 images [21, 22]. For instance, statistical similarities between morphological features of two brain regions, such as cortical thickness [23], gyrification index, surface area [24], and GMV [19], can be used to build a personalized morphological network. This network is then modeled as a graph for subsequent connectomic analysis. Unlike group-level structural covariance networks—which assess the correlation or covariance of brain structure across different brain regions in a population [25, 26]—SSMN emphasizes individual differences in morphological connectivity. The statistical interdependence between regions reflects the coordinated development or synchronized maturation of brain areas, influenced by the cortical microarchitecture, such as gene expression, cytoarchitectonic classification, and myelin content [21, 22, 27]. This personalized approach allows for a more nuanced exploration of individual variability in brain connectivity.

SSMN has demonstrated high robustness and reliability, and nontrivial topological properties [19], as well as neurobiological substrates [20, 27]. Moreover, it shows a strong resemblance to tractography- and fMRI-based connectome [28, 29]. Building on these methodological advancements, SSMN has been utilized to investigate normal brain development, aging, and a range of neuropsychiatric disorders [21, 22], including psychotic disorders [30], major depressive disorder [31], Alzheimer’s disease [32], autism spectrum disorder [33]. Alterations in SSMN observed in these populations have demonstrated strong correlations with, or predictive value for cognitive development and clinical outcomes. Despite these promising applications, the clinical significance of SSMN in understanding post-stroke deficits and recovery remains largely unexplored, highlighting the need for further investigation in this area.

In this study, we investigated the clinical relevance of SSMN in a sizable cohort of stroke patients who underwent longitudinal structural MRI imaging and comprehensive behavioral and cognitive assessments [2, 34]. The assessments spanned multiple domains, including motor skills, language, attention, and memory. Each patient’s SSMN was constructed using the Automated Anatomical Labeling (AAL) atlas, comprising 90 parcels, by evaluating dissimilarities in regional probability density functions derived from voxel-wise GMV vectors. For validation, SSMNs were also constructed using the Brainnetome atlas, which provides a finer-grained parcellation with 246 parcels [35]. Disruptions in edge-level and global topological properties of the SSMN during the acute and recovery phases were analyzed by correlating them with behavioral and cognitive deficits. We hypothesized that stroke-induced structural damage would manifest as a disconnectome detectable via the SSMN, characterized by altered network topology, lesion-dependent disruptions in morphological connectivity, and robust associations with behavioral and cognitive impairments. Furthermore, we anticipated that changes in the coupling between SSMN properties and behavioral/cognitive functions during recovery would reflect the dynamic adaptation of brain networks in response to stroke.

## Methods

We used a publicly available stroke functional neuroimaging dataset (https://cnda.wustl.edu/data/projects/CCIR_00299). The dataset includes longitudinal structural imaging, resting-state fMRI and neuropsychological test scores at 2 weeks, 3 months, and 1 year after stroke (Figure 1).

**Figure 1.**
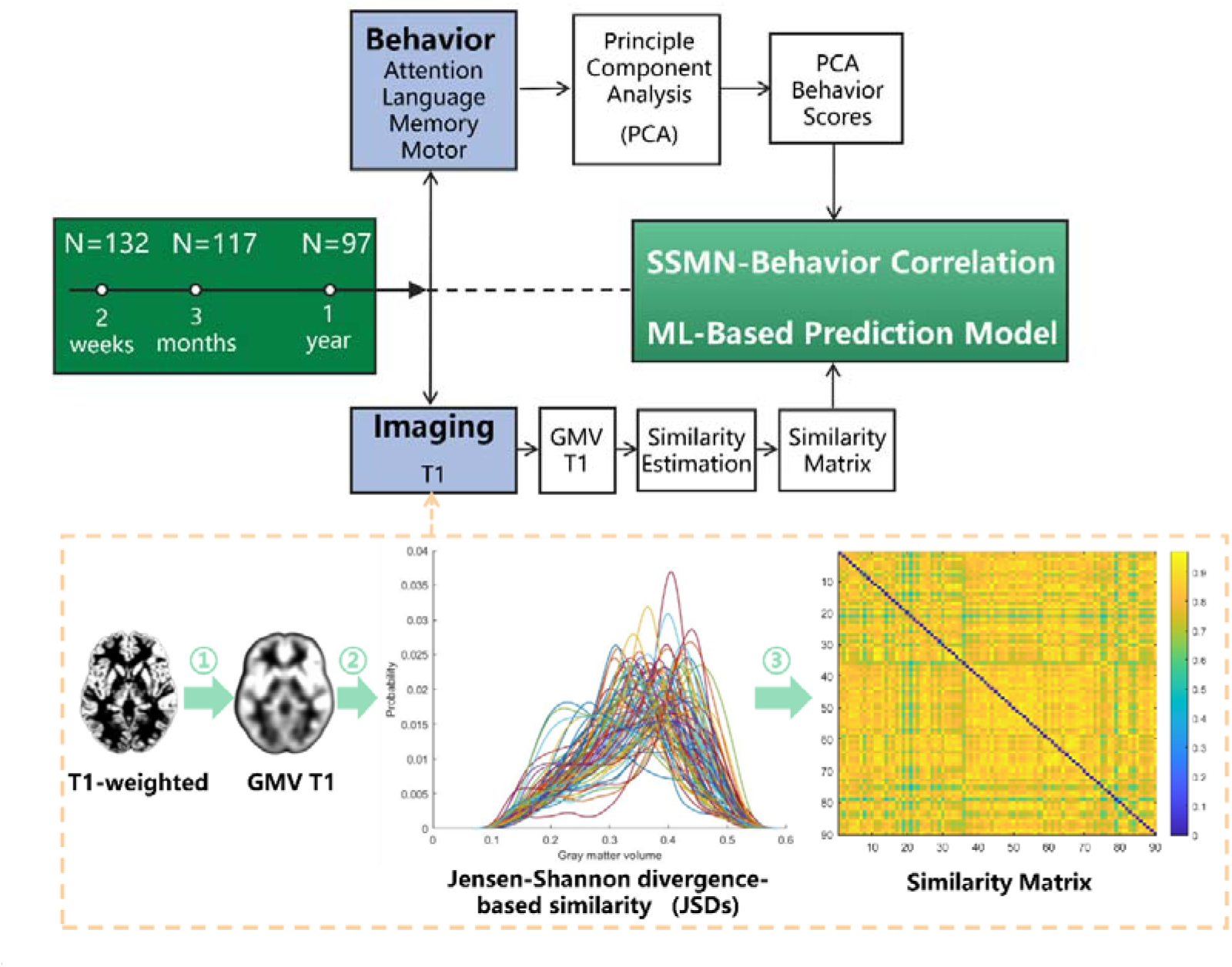
Schematic flowchart depicting the data acquisition, SSMN construction, and SSMN-behavior analysis. Longitudinal high-resolution T1w images and behavior data were acquired from a large cohort of patients after stroke. SSMN was constructed by evaluating dissimilarities among regional probability density functions derived from gray matter volume. We assessed the behavioral relevance of SSMN by assessing the SSMN-deficits correlation patterns and constructing machine-learning-based prediction models. GMV: Gary matter volume.

### Patients

The dataset included 132 patients with their first symptomatic stroke. Details of the data are described in Corbetta et al. [2]. As summarized in Corbetta et al [2], the inclusion criteria were: (1) age 18 years or older. There is no upper age limit. (2) First symptomatic stroke, ischaemic or haemorrhagic. (3) Up to two lacunes, clinically silent, less than 15 mm in size on CT scan. (4) Clinical evidence of motor, language, attention, visual, or memory deficits based on neurological examination. (5) Time of enrollment: within two weeks from stroke onset. (6) Awake, alert, and able to participate in the research.

Exclusion criteria. (1) Previous stroke based on clinical imaging. (2) Multifocal strokes. (3) Inability to remain awake during the study. (4) Presence of other neurological, psychiatric, or medical conditions that may preclude active participation in research and/or may alter the interpretation of the behavioral/imaging studies (e.g., dementia, schizophrenia) or limit life expectancy to less than one year (e.g., cancer or class IV heart failure). (5) Report of claustrophobia or metal object in the body.

### Healthy controls

Thirty-one healthy controls (HCs) were recruited. Exclusion criteria were: (1) a history of neurological, psychiatric, or medical abnormalities preventing participation in research activities, (2) a history of atherosclerotic (coronary, cerebral, peripheral) artery disease, or (3) an abnormal neurological examination with signs of central nervous system dysfunction.

### MRI imaging

MRI imaging data was acquired with a Siemens 3T Tim-Trio scanner at the School of Medicine of Washington University in St. Louis. Structural scans consisted of: (1) a sagittal MPRAGE T1-weighted image, TR=1950 msec, TE=2.26 msec, flip angle=9 deg, voxel size=1.0 x 1.0 x 1.0 mm, slice thickness = 1.00 mm; (2) a transverse turbo spin-echo T2-weighted image, TR=2500 msec, TE=435 msec, voxel-size=1.0 x 1.0 x 1.0 mm, slice thickness = 1.00 mm; and (3) a sagittal FLAIR, fluid attenuated inversion recovery, TR=7500 msec, TE=326 msec, voxel-size=1.5 x 1.5 x 1.5 mm, Slice thickness = 1.50 mm.

In this study, the T1-weighted images were used for SSMN, and the T2-weighted and FLAIR images were used for lesion drawing. The rs-fMRI data have been previously published to investigate functional disconnectome [7, 36–38].

### Neuropsychological assessment

All participants underwent behavioral tests including batteries of motor, language, attention, and memory. The behavioral tests were performed after scanning, and usually on the same day. Motor function tests included upper and lower body function, such as range of motion, grip strength, dexterity of hands, walking, and lower extremity strength. Language tests included word discrimination, naming, reading, and stem completion. Subtests of the Boston Diagnostic Aphasia Examination (BDAE-III) were used. Different visuospatial attention processes were measured using the Posner orienting task. Visuomotor spatial deficits were assessed with the Star Cancellation subtest of the Behavioral Inattention Test (BIT), and the Mesulam Unstructured Symbol Cancellation Test. Visual memory was studied with the Brief Visuospatial Memory Test-Revised (BVMT-R) (Benedict, 1997). Verbal memory was assessed with the Hopkins Verbal Learning Test-Revised (HVLT-R) (Brandt and Benedict, 2001). Spatial working memory was examined with a subtest of the Wechsler Memory Scale (Psychological Corp, 1981).

### Behavioral data reduction

Principal component analyses (PCA) were conducted for each domain. Components were retained if they met the following criteria: (1) eigenvalues greater than 1 and (2) accounted for more than 10% of the variance.

### 3D-T1 images processing

This study used CAT (Computational Anatomy Toolbox, http://www.neuro.uni-jena.de/cat) to calculate gray matter volume. In order to reduce the effect of head movement on structural MRI, two types of quality control were performed. All images were first checked for quality by visual and then quantified and graded by CAT for image resolution, noise and bias distortion. Images with a weighted average overall quality of less than 70% were excluded.

The VBM process is initiated by the application of a spatially adaptive non-local means (SANLM) denoising filter, followed by the application of SPM’s standard unified segmentation. The segmented files (i.e. grey matter, white matter and cerebrospinal fluid) serve as a starting point for further optimisations and CAT’s tissue segmentation steps. First, the brain is parcellated into left and right hemispheres, subcortical areas, ventricles and cerebellum. Second, a local intensity transformation is applied to reduce the effects of higher grey matter intensities in the motor cortex, basal ganglia and occipital lobe. Third, an adaptive maximum a posteriori (AMAP) segmentation is applied, which does not rely on a priori information about tissue probabilities. The AMAP segmentation also includes a partial volume estimation. Fourth, DARTEL (Diffeomorphic Anatomical Registration using Exponentiated Lie algebra) 26 is used to register the individual tissue segments to standardised templates in the ICBM 2009c Nonlinear Asymmetric space. Finally, modulation was applied to adjust for volume changes due to registration and spatial smoothing by convolution with a Gaussian kernel (FWHM = 8).

### SSMN

A graph or network consists of nodes (brain regions) and the edges connecting these nodes. In this study, we constructed large-scale morphological similarity networks, with nodes representing brain regions from the AAL atlas (90 parcels) and edges reflecting the interregional similarity in the distributions of regional gray matter volume (Figure 1). The interregional similarity was assessed with Jensen-Shannon divergence-based similarity (JSD). Given that different brain parcellation schemes lead to distinct topological organizations in brain networks [39–42], we also performed validation analyses using the human Brainnetome atlas (246 parcels) [35].

### Definition of network edges

To estimate SSMN between nodes, a Jensen – Shannon divergence-based similarity(JSDs) measure was adopted to quantify morphological connectivity between two regions [19] (Figure 1). JS divergence is an index from probability theory that measures the difference between two probability distributions or the information lost when a probability distribution is used to approximate another from the perspective of information theory. For each participant, the gray matter volumes for all voxels within a parcel were first extracted and used to estimate the kernel density estimation (KDE) with bandwidths chosen automatically. The probability distribution function (PDF) was calculated from the probability density function estimated using kernel density estimation for these values. JS divergence between the two parcels was estimated based on their PDFs. The formula for calculating the JSDs between points P and Q is as follows:

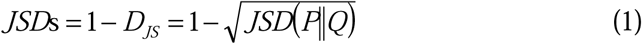

Where

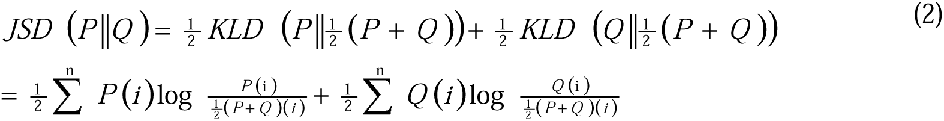

In mathematical statistics, the Kullback-Leibler Divergence (KLD) quantifies the difference between two probability distributions. The JSD ranges from 0 to 1, where 0 represents different distributions, and 1 represents identical distributions.

### Network analysis

For each of these networks, we computed both global measures (clustering coefficient *C_p_*, characteristic path length *L_p_*, local efficiency *E_loc_*, global efficiency *E_glob_*, and modularity *Q*) and nodal measures (nodal degree *k_i_*, nodal efficiency *e_i_*, and nodal betweenness *b_i_*). The clustering coefficient quantifies the extent of local clustering or cohesiveness within a network. The characteristic path length serves as an indicator of the network’s overall routing efficiency. Local efficiency assesses the capacity for parallel information propagation within localized subgraphs, as well as the network’s resilience to disruptions. Global efficiency evaluates the potential for parallel information transmission across the entire network. Modularity quantifies the degree to which nodes can be organized into subsets defined by dense internal connections and sparse external links. The nodal degree reflects the number of edges connected to a specific node within the network. Nodal efficiency measures the effectiveness of communication between a node and other nodes, while nodal betweenness quantifies a node’s influence on the flow of information among its peers.

### Disconnection and disconnectome for patients with similar stroke topology

To assess the disconnection and disconnectome by focal lesion, we selected patients with similar stroke lesion location. Due to the lesion heterogeneity (Figure 2), eleven and ten patients with circumscribed lesions in the left and right putamen were selected, respectively. The disconnection diaschisis was assessed by comparing the edge strength with HCs. The disconnectome was assessed by calculating the topological properties of small-worldness (sigma, lambda, and gamma) and comparing them with HCs. Before topological characterization of the SSMN matrices, a thresholding procedure is typically used to exclude noisy elements. Here, we employed a sparsity threshold, S (the ratio of actual edges divided by the maximum possible number of edges in a network), to convert each matrix into a binary network. The sparsity threshold method ensures the same number of nodes and edges for the resultant networks across participants. Due to the lack of a conclusive method to select a single threshold, a consecutive sparsity ranges from 0.02 to 0.4 with a 0.01 interval was chosen [19].

**Figure 2.**
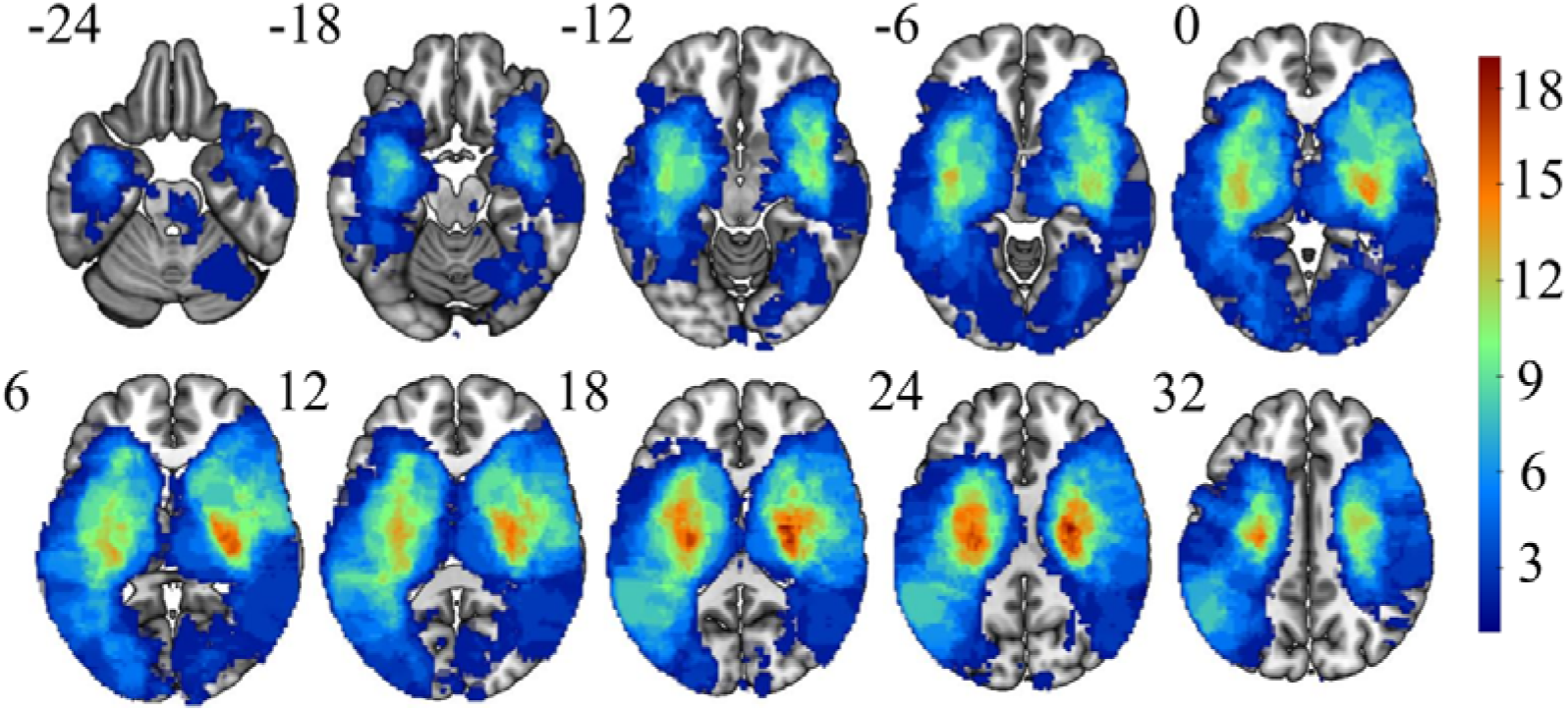
Lesion topography of the 103 patients. The color bar represents the number of patients with a lesion on a specific voxel.

Detailed introductions and interpretations and the mathematical definitions of these graph metrics can be seen in Rubinov and Sporns [43]. These analyses were performed using the GRETNA toolkit (https://www.nitrc.org/projects/gretna/) [44]

### Statistics analyses

Age and education were compared using two-sample t-tests. Sex difference was analyzed with Pearson’s chi-square test. The differences in deficits between HCs and patients, as well as their recovery, were assessed using a linear mixed-effect model (the *nlme* package in *R*).

Between-group differences in edge and global topological properties were analyzed using two-sample t-tests. For edge strength, results were corrected using network-based statistics (NBS) (Zalesky, Fornito, & Bullmore, 2010), with an edge *p*-value of 0.05 and a component *p*-value of 0.05. Global topological properties were corrected using False Discovery Rate (FDR), with a corrected *p*-value of 0.05.

The associations between SSMN and patients’ PCA scores were evaluated by calculating partial Pearson correlation coefficients, after accounting for covariates of non-interest. Details of these covariates are provided in the Supplementary table 1 for each domain. The results of the partial correlations were corrected using network-based statistics (NBS) with a component-level p-value threshold of 0.05.

### Machine learning-based SSMN-deficits prediction model

Apart from univariate analyses between SSMN and behavioral/cognition deficits, we also constructed multivariate machine learning-based SSMN-deficits prediction models. The relevance vector regression (RVR) algorithm and linear kernel function were adopted [45, 46]. RVR has no algorithm-specific parameter and thus does not require extra computational resources to estimate the optimal algorithm-specific parameters [47].

#### Linear relevance vector regression (RVR)

RVR is a Bayesian framework for learning sparse regression models. In RVR, only some samples (smaller than the training sample size), termed the ‘relevance vectors’, are used to fit the model:

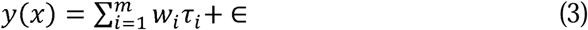

where *τ_i_* are basis functions, ∈ is normally distributed with mean 0 and variance *β*. RVR uses training data to build a regression model:

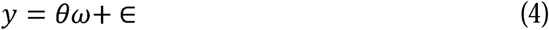

where 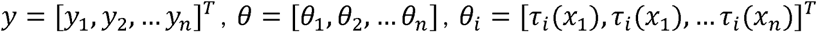. Each vector *θ_i_*, consisting of the values of the basis function *τ_i_* for the input vectors, is a relevance vector.

The model parameters *β* were found by using the maximum likelihood estimates from the conditional distribution: 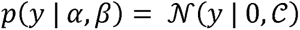, where the 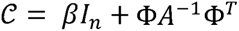. To make the RVM favor sparse regression models, prior distributions were assumed for both *w_i_* and 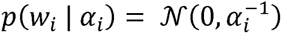. The ways of treating priors, however, lead to the same relevance vector machine construction.

#### Prediction accuracy and significance

Leave-one-out cross-validation (LOOCV) was used to calculate the prediction accuracy (the Pearson correlation coefficient between the predicted and actual labels). In each turn of the LOOCV, one patient was designated as the test sample, and the remaining patients were used to train the lesion model. The predicted score was then obtained by the feature matrix of the tested sample.

The significance level was computed based on 1000 permutation tests. The prediction labels (e.g., PCA scores of language function) were randomized for each permutation test, and the same RVR prediction process used in the actual data was carried out. After 1000 permutations, a random distribution of accuracies was obtained and the *P* value was correspondingly calculated: 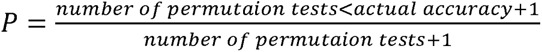.

The features of each prediction model were the subject’s SSMN matrix.

## Results

### Stroke lesion topography

Due to severe head movement and poor overall quality of T1-weighted images, 29 patients were excluded from the analysis. Consequently, 103 patients were included in the SSMN analyses. Additionally, 28 age- and sex-matched HCs were selected for between-group comparisons. The sample sizes varied across different time points and domains, with a maximum of 103 patients available for language assessments two weeks post-stroke (Table 1).

**Table 1.** Demographic and clinical information of healthy controls and stroke patients.

| Variables | Controls(n=28) | Stroke(n=103) | *P* value |
| --- | --- | --- | --- |
| Age | 54.29±11.34<br>(21~79) | 52.3 ± 11<br>(27~72) | 0.394 |
| Gender(male/female) | 13/15 | 56/47 | 0.46 |
| Lesion_type |  |  | - |
| Ischemic | - | 80(78%) |  |
| Hemorrhagic | - | 16(15%) |  |
| Other |  | 7(7%) |  |
| Race |  |  | 0.94 |
| White | 10(36%) | 36(35%) |  |
| Black or African American | 18(64%) | 67(65%) |  |
| Years of education | 13(12~21) | 13(9~20) | 0.786 |
| Smoke | 12(43%) | 52(51%) | 0.474 |
| High blood pressure | 0(0%) | 68(66%) | <0.001 |
| Diabetes | 4(14%) | 27(26%) | 0.272 |

Gender, smoking status, high blood pressure, and diabetes were assessed using Chi-squared tests, while age and education levels were evaluated using the two-sample t-tests.

The lesion locations of the 103 patients were predominantly distributed bilaterally in the white matter and subcortical regions. Cortical lesions were primarily localized within the territory of the middle cerebral artery, affecting the frontal, parietal, and temporal lobes (Figure 2).

### PCA-based behavioral and cognitive deficits and recovery

PCA results for each domain, as reported by Corbetta et al. [2], were publicly available and are summarized in Figure 3, which illustrates post-stroke deficits and recovery trajectories. To evaluate recovery patterns, we performed linear mixed-effects model analyses, incorporating group and post-stroke time as fixed effects and subjects as random effects. The analyses revealed significant main effects of group and post-stroke time across multiple domains (Supplementary Table 2), including language (*Ps* < 0.001), left motor function (*Ps* < 0.001), right motor function (*Ps* < 0.001), spatial memory (*Ps* < 0.001), verbal memory (*Ps* < 0.001), and average performance attention (group effect: *P* = 0.009; post-stroke time effect: *P* = 0.026). For validity/disengagement attention, a significant group effect (*P* = 0.0334) and a marginal post-stroke time effect (*P* = 0.061) were observed. Notably, significant interaction effects between group and post-stroke time were identified for language (*P* = 0.0037), spatial memory (*P* = 0.0031), and average performance attention (*P* < 0.001), underscoring the dynamic interplay of these factors over time. These findings highlight the complexity of recovery trajectories across different cognitive and behavioral domains.

**Figure 3.**
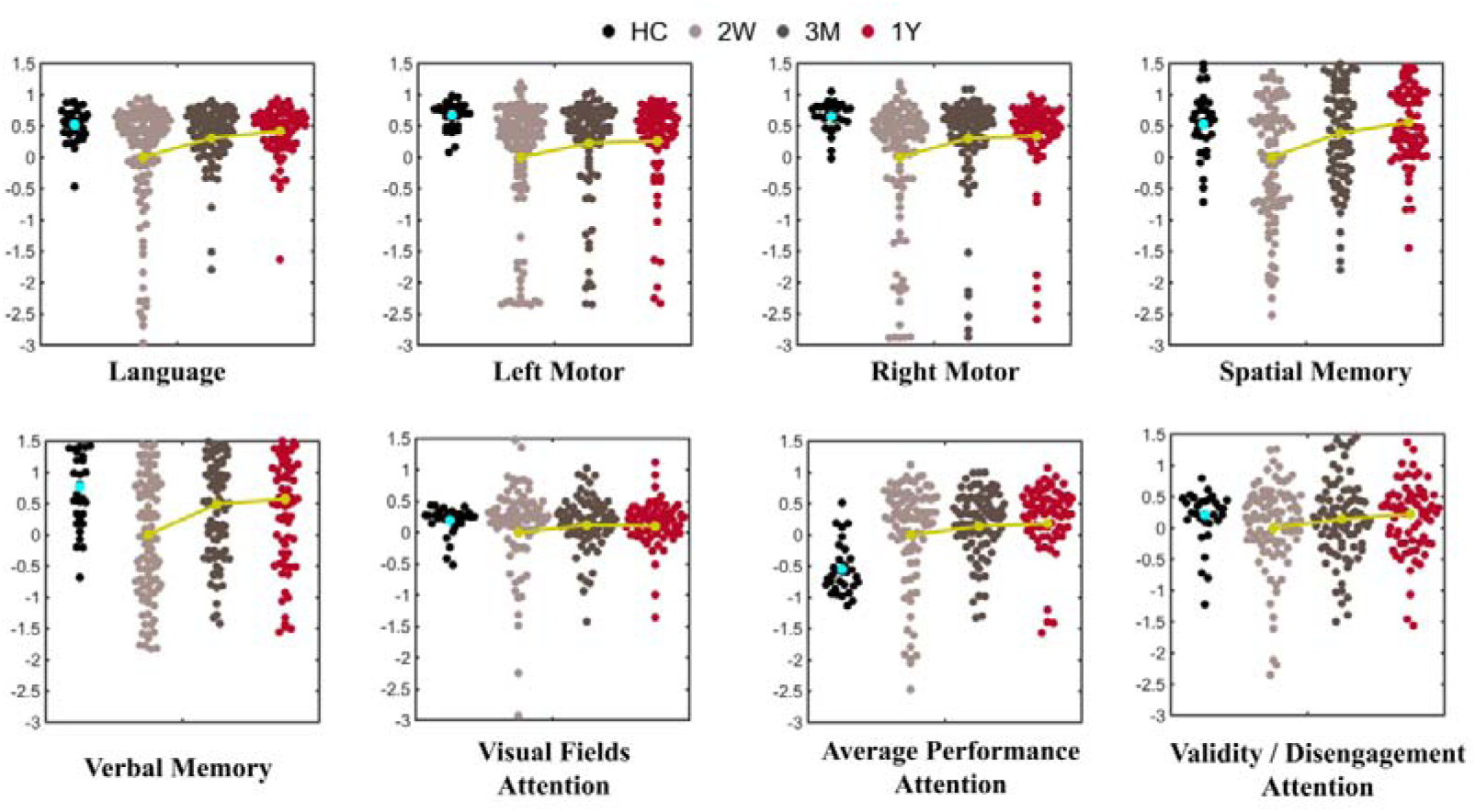
Behavioral and cognitive deficits for the eight domains.

### Disconnection and disconnectome at two weeks after subcortical stroke

For patients who experienced a stroke in either the left or right putamen, lesion location-dependent disconnection patterns were observed (Figure 4). Strokes in the left putamen resulted in network disruptions involving the left superior temporal gyrus (STG), inferior frontal gyrus (IFG), rolandic operculum, supramarginal gyrus (SMG), anterior cingulate cortex (ACC), postcentral gyrus (PoCG), and the right hippocampus. In contrast, strokes in the right putamen caused network disruptions in the right amygdala, thalamus, rolandic operculum, putamen, hippocampus, and STG, as well as the left Heschl’s gyrus, ACC, and insula.

**Figure 4.**
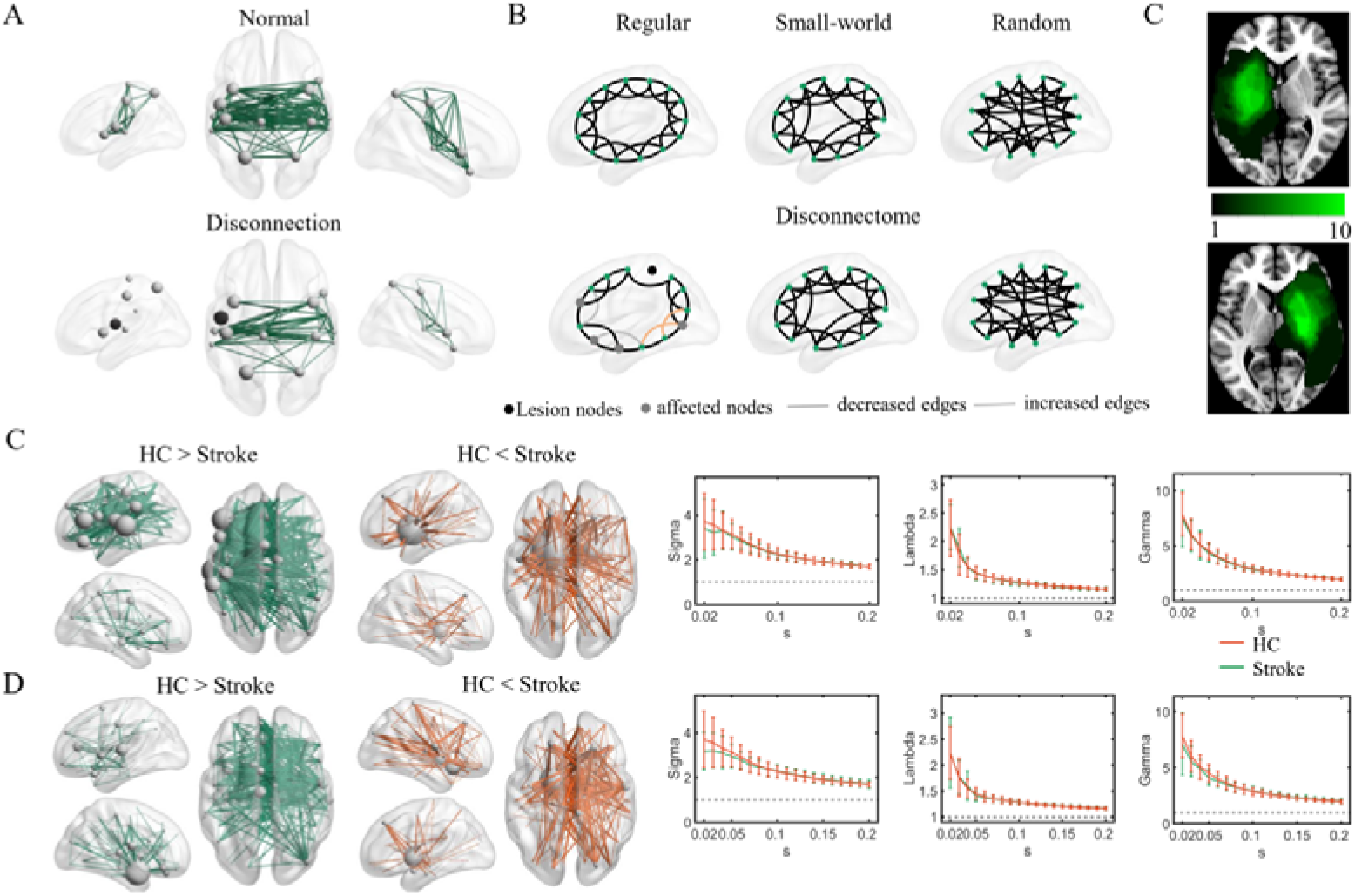
Disconnection and Disconnectome assessed by SSMN. A&B: A schematic diagram illustrating disconnection and disconnectome. In disconnection, a focal lesion induces local and distant network disruptions. In disconnectome disruption, a focal lesion compromised network topology. B: Lesion topography patients with circumscribed strokes in the left putamen (n = 11) and the right putamen (n = 11). C&D: Empirical evidence of disconnection and disconnectome in the two subgroups assessed by SSMN. Lesion-location-dependent SSMN disruptions were observed when compared to HCs. For connectomal analyses, SSMN demonstrated small-worldness for HCs and patients, but no significant difference was observed.

In both subgroups, hyper-morphological connections were identified, with the highest connectivity localized in nodes situated within or near the lesion topography. These hyper-morphological connections may be associated with increased gray matter volumes (GMVs) in the stroke lesion areas. To evaluate GMV changes within the stroke lesion, we compared the GMVs in these regions to the total GMV of the corresponding homotopic areas. Supplementary Figure 1A illustrates an example of a patient demonstrating hyper GMV within the stroke lesion. Statistical analyses revealed significantly elevated GMVs within stroke lesions compared to homotopic regions at both 2 weeks and 3 months post-stroke (*Ps* < 0.001; Supplementary Figure 1B). By 1 year post-stroke, these differences were no longer significant, indicating a normalization of GMV within the stroke lesion over time.

No significant differences in small-worldness were identified between patients and healthy controls (Figure 4). However, when the entire sample (n = 103) was considered, a decrease in Lambda was observed, suggesting a transition to a more regular but less efficient network configuration (Supplementary Figure 2).

### Domain-specific correlation patterns between SSMN and post-stroke deficits

For the SSMN connections associated with post-stroke deficits, we investigated whether the brain regions they connected aligned with the neurobiological underpinnings of each domain or the correlation patterns established in prior studies using other imaging modalities, such as resting-state functional connectivity (rsFC) and voxel-based lesion-symptom mapping.

In the language domain, at two weeks post-stroke, left intrahemispheric (156 out of 406) and interhemispheric (211 out of 406) connections among the inferior frontal gyrus (IFG), superior temporal gyrus (STG), middle temporal gyrus (MTG), angular gyrus (AG), and primary motor cortices were significantly and positively correlated with language deficit scores (Figure 5, Supplementary Figure 3). These findings were consistent with the left-lateralized neurobiology of language and prior rsFC studies (Supplementary Figure 4).

**Figure 5.**
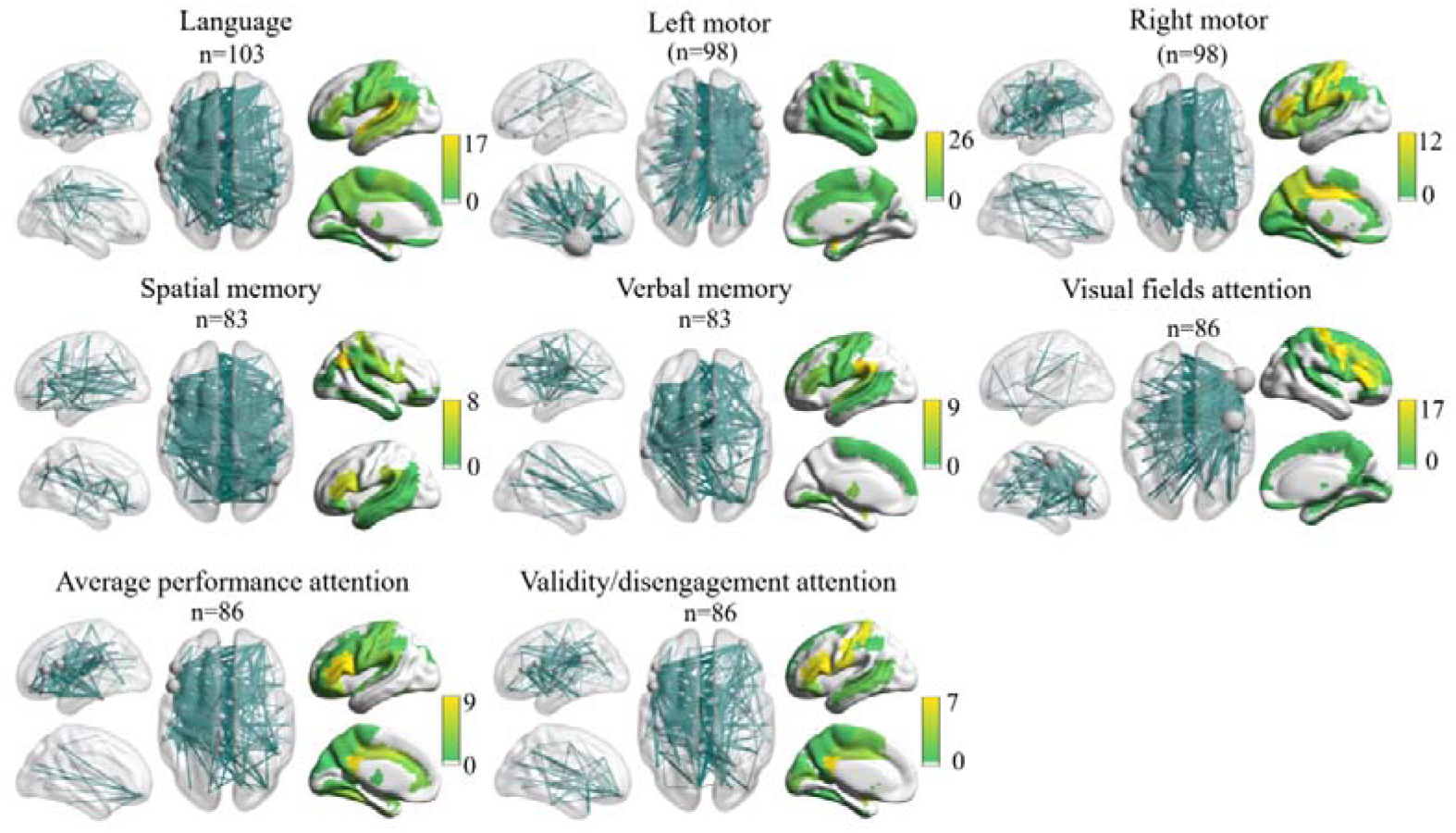
SSMN-deficits positive correlations at two weeks after stroke. Left-lateralized correlation patterns were observed for deficits across multiple domains, including language, right motor, verbal memory, average performance attention, and validity/disengagement attention. A right-lateralized correlation pattern was observed for deficits in the left motor. Edge *p*<.05, component *p*<.05 with NBS correction.

Motor deficits exhibited distinct lateralized patterns of network disruption. Left motor deficits were strongly associated with right-lateralized network disruptions, involving intrahemispheric (111 out of 400) and interhemispheric (276 out of 400) connections among the right prefrontal cortex, postcentral gyrus, STG, supramarginal gyrus (SMG), and the left insula, middle frontal gyrus (MFG), and dorsal parietal cortex. Conversely, right motor deficits were linked to left-lateralized network disruptions, with significant intrahemispheric (139 out of 378) and interhemispheric (198 out of 378) connections among the left primary motor cortex, IFG, and insula, as well as the right precentral gyrus, superior frontal gyrus (SFG), and dorsal parietal cortex, correlating positively with right motor deficit scores.

In the domain of spatial memory, significant positive correlations were observed between spatial memory deficit scores and connections involving the bilateral inferior frontal gyrus (IFG), supramarginal gyrus (SMG), superior temporal gyrus (STG), right angular gyrus (AG), postcentral gyrus, and STG. For verbal memory deficits, significant positive correlations were identified in 145 connections, including left intrahemispheric connections (52 out of 145) and interhemispheric connections (71 out of 145) involving the left SMG, IFG, STG, middle temporal gyrus (MTG), and primary motor cortex.

Attention deficits exhibited distinct lateralized patterns of network disruption based on the subdomain. Visual field attention deficits were associated with strong right-lateralized network disruptions, while average performance attention and validity/disengagement attention deficits were linked to strong left-lateralized network disruptions. Visual Field Attention Deficits: Significant positive correlations were identified in 232 connections, comprising 80 right intrahemispheric and 104 interhemispheric connections. These involved the right precentral gyrus, IFG, middle frontal gyrus (MFG), STG, and left insula, medial prefrontal cortex, and parietal cortices. Average Performance Attention Deficits: Significant correlations were observed in 244 connections, including 104 left intrahemispheric and 127 interhemispheric connections. These involved the left IFG, posterior cingulate cortex (PCC), midcingulate gyrus (MCG), and primary motor cortex. Validity/Disengagement Attention Deficits: Significant positive correlations were noted in 211 connections, with 70 left intrahemispheric and 104 interhemispheric connections. These connections involved the left IFG, PCC, postcentral gyrus, MTG, and the right precentral gyrus, MCG, and PCC.

At three months and one year post-stroke, significant positive correlations between SSMN connections and post-stroke deficits were also observed (Supplementary Figures 5, 6, 8, and 9). Compared to the patterns identified at two weeks post-stroke, brain regions associated with the neurobiological mechanisms of different domains maintained significant correlations, and the overall lateralization pattern remained consistent. However, the number of positively correlated connections decreased substantially, suggesting that morphological networks undergo significant reorganization as symptoms improve (Supplementary Figures 3, 7, and 10).

### SSMN connections implicated across multiple domains

To determine whether certain SSMN connections were implicated across multiple domains, we calculated the sum of positive correlation values across all domains in each time (Supplementary Figure 11). At two weeks post-stroke, the regions exhibiting the highest correlations were the left inferior frontal gyrus (IFG), cingulate gyrus, hippocampus, and supramarginal gyrus (SMG). By three months post-stroke, the highest correlations were observed in the left medial orbitofrontal cortex. At one year post-stroke, the region with the highest correlation shifted to the right inferior temporal gyrus (ITG).

### Correlation patterns between post-stroke SSMN changes and behavioral and cognitive recovery

Time- and domain-specific correlation patterns between SSMN changes and behavioral and cognitive recovery were identified (Figures 6 and 7). Notably, language recovery exhibited distinct associations with specific brain regions in the right hemisphere across different recovery phases. Between two weeks and three months post-stroke, language recovery was significantly correlated with SSMN changes in the right precentral gyrus. However, from three months to one year post-stroke, the primary correlation shifted, with language recovery becoming predominantly associated with SSMN changes in the right temporal cortex.

**Figure 6.**
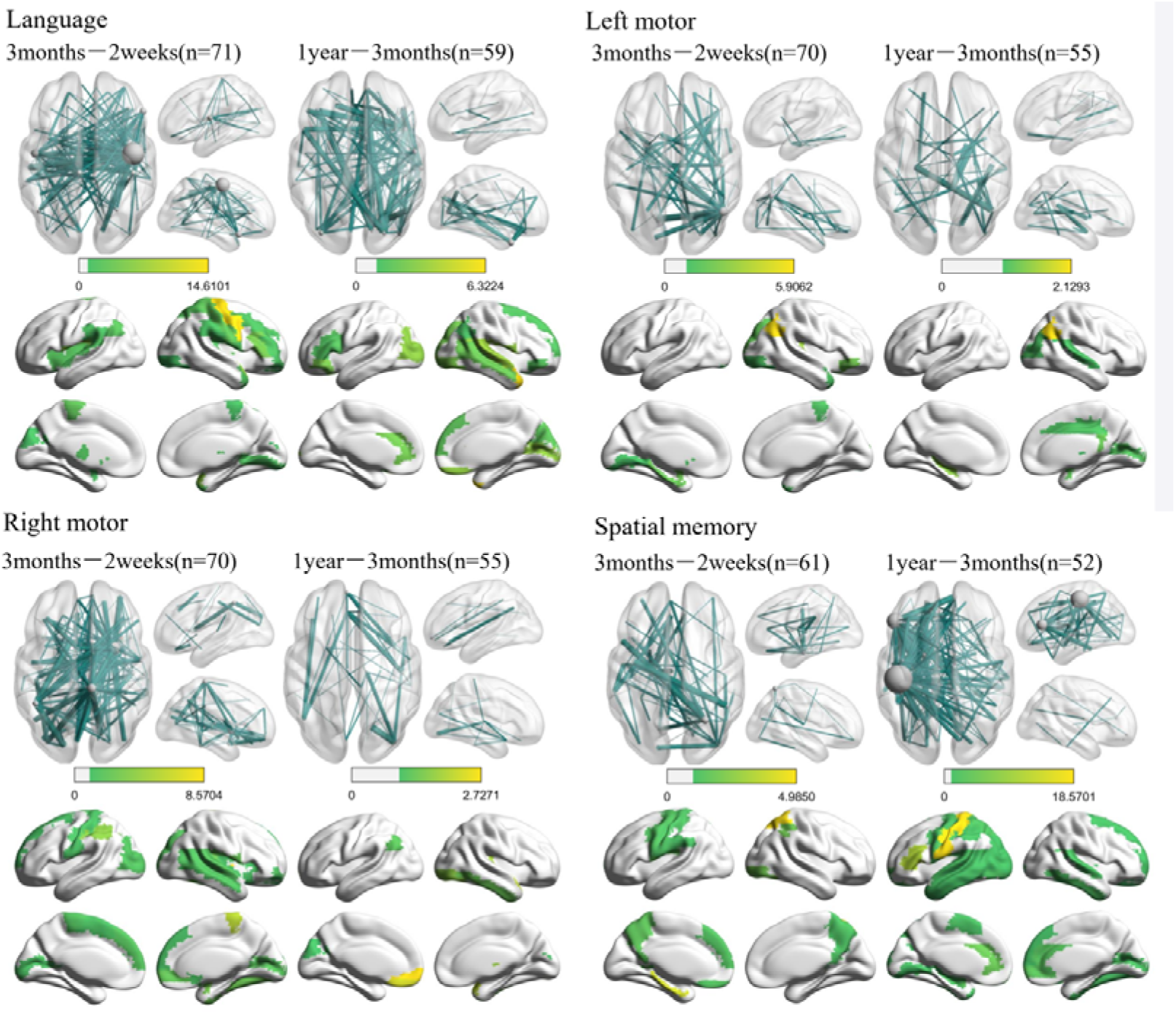
The Pearson correlation patterns between SSMN normalization and behavioral/cognitive recovery for domains of language, left and right motor, and spatial memory. Edge *p*<.05, component *p*<.05 with NBS correction.

**Figure 7.**
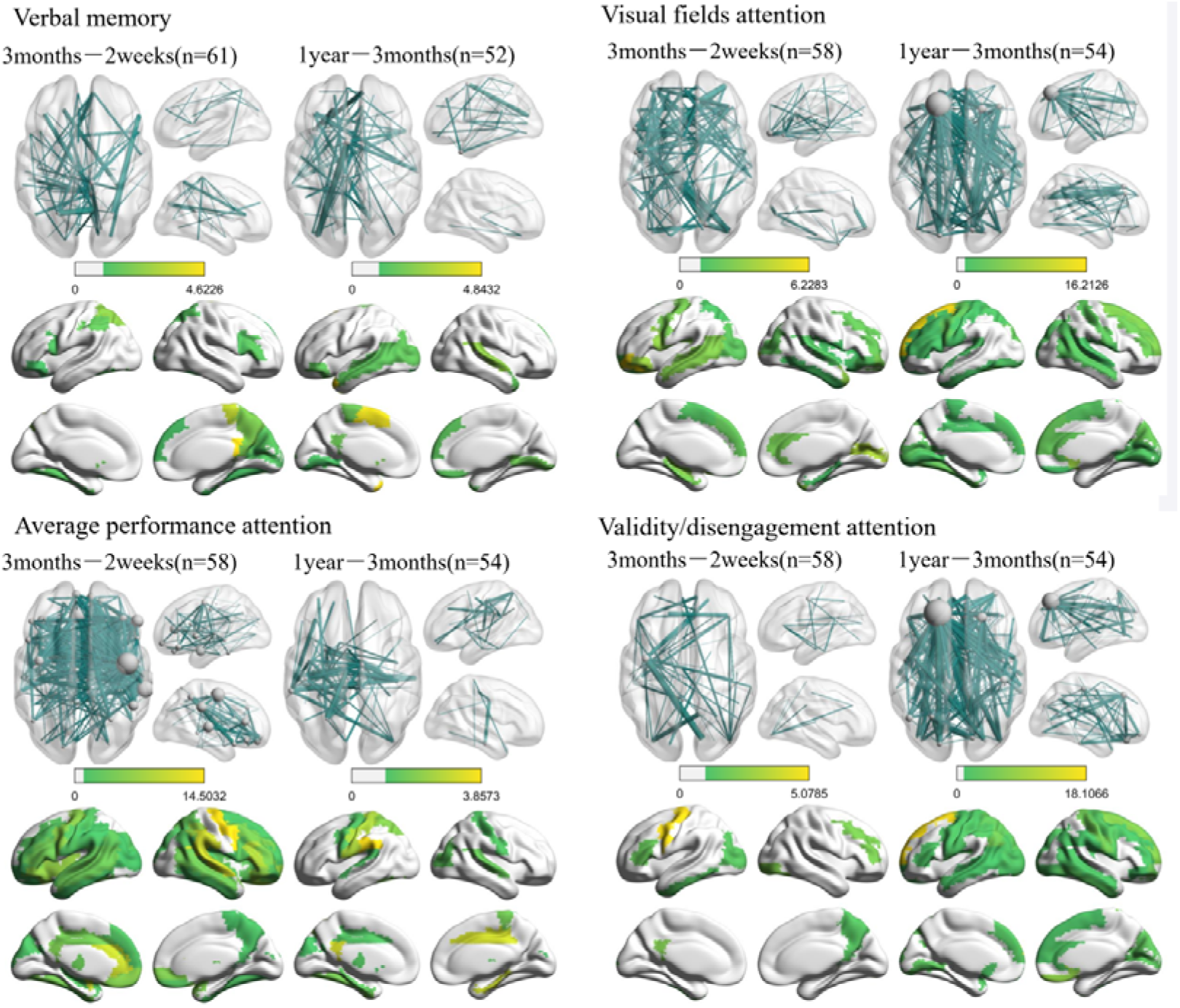
The Pearson correlation patterns between SSMN normalization and behavioral/cognitive recovery for domains of verbal memory, visual field attention, average performance attention, and validity/disengagement attention. Edge *p*<.05, component *p*<.05 with NBS correction.

### Results of SSMN-deficits prediction

RVR-based SSMN-deficits prediction analyses revealed that SSMNs were significant predictors of post-stroke behavioral and cognitive deficits (Table 2 and Supplementary Figures 12, 13, and 14): language (at two weeks: *R* = 0.46, permutation *P* < 0.001; at three months: *R* = 0.275, permutation *P* = 0.025; at one year: *R* = 0.488, permutation *P* = 0.002), left motor (at two weeks: *R* = 0.642, permutation *P* < 0.001; at three months: *R* = 0.332, permutation *P* = 0.022; at one year: *R* = 0.655, permutation *P* < 0.001), right motor (at two weeks: *R* = 0.473, permutation *P* < 0.001; at three months: *R* = 0.328, permutation *P* =0.015; at one year: *R* = 0.325, permutation *P* = 0.015), spatial memory (at two weeks: *R* = 0.53, permutation *P* < 0.001; at three months: *R* = 0.257, permutation *P* = 0.128; at one year: *R* = 0.329, permutation *P* = 0.012), verbal memory (at two weeks: *R* = 0.36, permutation *P* < 0.001; at three months: *R* = 0.006, permutation *P* = 0.466; at one year: *R* = 0.266, permutation *P* = 0.054), visual fields attention (at two weeks: *R* = 0.399, permutation *P* < 0.001; at three months: *R* = 0.203, permutation *P* = 0.108; at one year: *R* = 0.151, permutation *P* = 0.042), average performance attention (at two weeks: *R* = 0.264, permutation *P* < 0.001; at three months: cannot predict; at one year: *R* = 0.245, permutation *P* = 0.063), and validity/disengagement attention reached marginal significance (at two weeks: *R* = 0.214, permutation *P* = 0.075; at three months: *R* = 0.432, permutation *P* = 0.005; at one year: *R* = 0.172, permutation *P* = 0.127).

**Table 2.** RVR-based SSMN-deficits prediction results.

| Domain | 2 weeks |  | 3 months |  | 1 year |  |
| --- | --- | --- | --- | --- | --- | --- |
|  | R<br>(Permutation<br>*P*) | MAE | R<br>(Permutation<br>*P*) | MAE | R<br>(Permutation<br>*P*) | MAE |
| Language | **0.46**<br>(**< 0.001**) | 0.628 | **0.275**<br>(**0.025**) | 0.662 | **0.488**<br>(**0.002**) | 0.61 |
| Left motor | **0.642**<br>(**< 0.001**) | 0.584 | **0.332**<br>(**0.022**) | 0.767 | **0.655**<br>(**< 0.001**) | 0.539 |
| Right motor | **0.473**<br>(**< 0.001**) | 0.653 | **0.328**<br>(**0.015**) | 0.642 | **0.325**<br>(**0.015**) | 0.665 |
| Spatial memory | **0.53**<br>(**< 0.001**) | 0.663 | 0.257<br>(0.128) | 0.854 | **0.329**<br>(**0.012**) | 0.757 |
| Verbal memory | **0.36**<br>(**0.033**) | 0.796 | 0.006<br>(0.466) | 0.992 | 0.266<br>(0.054) | 0.8 |
| Visual fields<br>attention | **0.399**<br>(**0.002**) | 0.556 | 0.203<br>(0.108) | 0.706 | 0.151<br>(0.042) | 0.556 |
| Average performance<br>attention | **0.264**<br>(**0.037**) | 0.753 | -<br>(-) | 0.819 | 0.245<br>(0.063) | 0.689 |
| Validity/disengagement<br>attention | 0.214<br>(0.075) | 0.745 | **0.432**<br>**(0.005)** | 0.705 | 0.172<br>(0.127) | 0.764 |

### Validation results using the Brainnetome atlas

Prediction analyses confirmed that SSMN constructed using the Brainnetome atlas also significantly predicted deficits at two weeks post-stroke (language: *R* = 0.54, permutation *P* < 0.001; left motor: *R* = 0.59, permutation *P* < 0.001; right motor: *R* = 0.48, permutation *P* < 0.001; spatial memory: *R* = 0.32, permutation *P* = 0.012; verbal memory: *R* = 0.27, permutation *P* = 0.023), three months post-stroke (left motor: *R* = 0.299, permutation *P* = 0.02; right motor: *R* = 0.223, permutation *P* =0.05), one-year post-stroke (language: *R* = 0.26, permutation *P* = 0.038; left motor: *R* = 0.56, permutation *P* < 0.001; right motor: *R* = 0.26, permutation *P* =0.033; spatial memory: *R* = 0.33, permutation *P* = 0.016). See supplementary Table 3 and Supplementary Figures 15, 16, and 17 for details.

## Discussion

This study provides the first evidence that individualized morphological connectomes constructed using T1-weighted imaging hold clinical significance in understanding post-stroke disconnection and the resulting disconnectome. Post-stroke focal lesions disrupted both ipsilesional and interhemispheric morphological connections, with these disruptions demonstrating significant positive correlations with behavioral and cognitive deficits. Importantly, the correlation patterns observed within each domain were closely aligned with established neurobiological underpinnings and mirrored patterns identified through functional MRI and lesion-deficit mapping approaches. Additionally, we revealed that changes in SSMNs were closely associated with post-stroke behavioral and cognitive recovery, highlighting their potential as biomarkers for tracking rehabilitation progress. These findings underscore the utility of T1-weighted imaging as a novel modality for constructing and understanding the structural disconnectome following stroke.

Stroke syndromes result from widespread functional and structural disruptions in the brain. Understanding the disconnectome and its relationship with behavioral and cognitive deficits remains a central goal in clinical neurology. Functional MRI (fMRI) and diffusion tensor imaging (DTI) are currently the primary modalities used to map the human connectome in vivo. Despite its routine use in clinical practice, T1-weighted imaging has traditionally been viewed as a diagnostic tool aimed at identifying structural abnormalities in brain tissue. However, its potential for providing detailed morphological and connectivity insights has largely been overlooked. Compared to fMRI and DTI, T1-weighted imaging offers several advantages: it is faster and easier to acquire, relatively low-cost, widely available, and provides higher spatial resolution and signal-to-noise ratio. Recent studies have demonstrated the robustness, reproducibility, and reliability of sensorimotor structural networks (SSMNs) derived from T1-weighted imaging [19, 20, 23, 48]. In addition to its methodological advantages, the utility of SSMN in elucidating brain disconnection and the disconnectome is rooted in its neurobiological significance in constructing the human connectome. Morphological similarities between brain regions are thought to reflect coordinated developmental processes or synchronized maturation, underscoring their relevance in connectomics [49]. Growing evidence highlights that SSMNs capture key cellular, molecular, and functional characteristics of the brain [20, 27, 50]. Similar to the functional and tract-based structural connectomes, the morphological connectome exhibits intricate topological properties, including modular organization, small-world architecture, and rich-club phenomena [19, 50].

In this study, we established a robust relationship between SSMN disruptions and post-stroke behavioral and cognitive deficits, which aligned closely with the neurobiology of each domain. These correlation patterns also strikingly similar to findings from resting-state fMRI [7] and lesion-deficit mapping [51]. The resting fMRI of the patients used in this study, previously analyzed by Siegel et al. [7], revealed canonical functional connections and areas involved in each domain (Supplementary Figure 9). Similarly, anatomical damage associated with these deficits encompassed canonical cortical regions and their corresponding subcortical white matter tracts. This spatial alignment across modalities highlights the behavioral and cognitive relevance of SSMN. Furthermore, we compared the predictive performance of three neuroimaging metrics— resting-state brain networks, SSMN, and lesion territory— in predicting post-stroke symptoms (Supplementary Table 4). The maximum explained variance (*r^2^*) was 0.65 for lesion territory (language), 0.51 for resting-state brain networks (language), and 0.41 for SSMN (left motor). Although SSMN accounted for slightly less variance in post-stroke behavioral and cognitive impairments compared to the other two modalities, its alignment with established neuroimaging findings underscores its potential as a complementary tool for post-stroke assessment.

We further demonstrated that connectomal diaschisis following stroke is driven by disruptions in both ipsilesional and interhemispheric morphological connections, aligning with findings from fMRI [7] and DTI studies [52, 53]. Notably, interhemispheric disruption emerges as a pivotal feature of disconnectome after focal brain damages, and has been reported in a range of neurological disorders [54], including glioma [46, 55–57], multiple sclerosis [58], traumatic brain injury [59], and epilepsy [60]. The neurophysiological mechanisms underlying interhemispheric diaschisis remain poorly understood; however, research underscores the importance of homotopic connections and interhemispheric communication in supporting human cognition. Bilateral representation is particularly critical for functions such as attention, memory, and even language [61–63]. These findings highlight the intricate role of interhemispheric connectivity in maintaining functional integrity across the brain and its vulnerability in the context of stroke and other neurological conditions.

We did not observe a disconnectome in the two groups with circumscribed strokes localized to the putamen. This absence may be partially attributed to the limited sample sizes and the predominantly subcortical nature of the lesions in these patients, with relatively sparse cortical involvement. Notably, across the entire sample, less than 20% of cases involved cortical-only lesions. When comparing the full sample of stroke patients to healthy controls, we observed a significant increase in lambda values, but only at specific network thresholds (Supplementary Figure 2). This finding suggests the presence of connectomal diaschisis, characterized by a shift in network topology towards a more random organization. Such alterations in network topology are consistent with morphological studies in Alzheimer’s disease [64], depression [65], ADHD [66], ASD [67], and PTSD [68]. These results underscore the sensitivity of SSMN to detect network topological changes across various neurological and psychiatric disorders [21, 22], further highlighting its utility as a robust neuroimaging metric.

A critical mechanistic question is how stroke leads to the disruption of the SSMN. Insights from animal models of stroke have highlighted acute-phase processes such as cell death and axonal disruption [69, 70], as well as mechanisms of neural repair during recovery [71]. However, the link between macroscopic morphological changes and the underlying microscopic cellular and axonal alterations remains insufficiently explored. Notably, a rodent study provided compelling evidence of macro-micro coupling by comparing gray matter volume changes with histological markers of hippocampal damage in the same brains [72]. Following cardiac arrest and cardiopulmonary resuscitation, the CA1 region of the hippocampus exhibited significant neuronal loss and microglial invasion. Corresponding reductions in gray matter concentration were observed using T2-weighted imaging and were positively correlated with neuronal density and negatively correlated with microglial density in the CA1 region. This macro-micro relationship, along with the diffuse effects of stroke, suggests that stroke-induced macroscopic morphological changes in specific brain regions may underlie the disruption of the SSMN. During the recovery phase, morphological changes in affected brain regions may lead to adaptive remodeling of the SSMN. These changes in the SSMN could align with improvements in behavioral and cognitive functions, offering a dynamic biomarker for tracking the progression of stroke recovery.

There are several limitations in this study. First, due to the nature of stroke, the lesion territories were predominantly subcortical. Among patients with putamen stroke, white matter lesions were predominant, and no evidence of connectomal diaschisis was observed. However, it would be premature to conclude that SSMN is insensitive to connectomal diaschisis. Future studies focusing on samples with more circumscribed cortical lesions are needed to comprehensively assess connectomal diaschisis following stroke. Second, constructing personalized symptom prediction and rehabilitation models is critical for designing targeted interventions and treatments after stroke. Our findings demonstrate that SSMN significantly predicts deficits across different domains, with predictive performance comparable to other imaging modalities [2, 7]. However, SSMN considers only interconnections within gray matter. Since many stroke patients primarily experience subcortical damage, future research must address the unique contributions of distinct lesion types and tissues to develop more precise prediction models [73, 74]. Third, in this study, SSMN was constructed based on regional gray matter distribution similarity. However, gray matter morphology encompasses diverse characteristics, such as cortical thickness [23]. SSMN construction can leverage multiple features [21, 27], and employ various metrics to assess similarity among these features [22]. Recently, voxel-level SSMN construction has also been explored [20]. Although systematic evaluations of the comparative advantages and efficiency of these methodologies are currently lacking, other methods for constructing SSMN are also worth exploring.

## Conclusions

This study presents the first evidence that individualized morphological networks are sensitive to detecting disconnection and the disconnectome following stroke. Stroke-induced disruptions in ipsilesional and interhemispheric morphological connections were significantly associated with post-stroke behavioral and cognitive deficits. Furthermore, normalization of the SSMN was linked to recovery, highlighting its dynamic reorganization during recovery. These findings suggest that T1-weighted imaging offers a novel and valuable modality for analyzing the disconnectome in stroke patients.

## Supporting information

Supplementary results

## Data Availability

The datasets analysed during the current study are available in the https://cnda.wustl.edu/data/projects/CCIR_00299.

## Ethics approval and consent to participate

The data were part of the Washington University Stroke cohort. Written informed consent was obtained from all participants in accordance with the Declaration of Helsinki and procedures established by the Institutional Review Board of Washington University in Saint Louis. All participants were compensated for their time.

## Consent for publication

Not applicable

## Authors’ contributions

Binke Yuan: formal analysis, writing – original draft, funding acquisition, visualization;

Tao Zhong and Yaling Wang: formal analysis, writing – original draft, visualization;

Qingwen Chen, Xiaolin Guo, Junjie Yang, Xiaowei Gao, Zhe Hu, Junjing Li, Jiaxuan Liu, Zhiheng Qu, Wanchun Li, Zhongqi Li, Wanjing Li, Yien Huang, Jiali Chen, Hao Wen, Ye Zhang, Junle Li: writing – original draft

Han Gao: methodology, supervision, project administration, funding acquisition, writing – original draft.

## Acknowledgements

Not applicable

