## Supplementary results for "Using single-subject morphological networks to elucidate the patterns of disconnection and disconnectome associated with post-stroke deficits and recovery"

Supplementary Table 1. Covariates of non-interest accounted for SSMN-deficits correlation analysis.

| Domain | Two weeks | Three months | One year |
| --- | --- | --- | --- |
| Language | Age, Gender, TIV, Motor L, Attention (Vf, Ave, Val/Dis) | Age, Gender, Education, TIV, Motor (L, R), Attention (Vf, Ave, Val/Dis), Memory (S, V) | Age, Gender, Lesion size, TIV, Motor (L, R), Attention (Vf, Ave, Val/Dis |
| Left Motor | Age, Education, Lan, Motor R, Attention Val/Dis, Memory V | Age, Gender, Education, TIV, Language, Motor R, Attention (Ave, Val/Dis), Memory (S, V) | Age, Gender, TIV, Language, Motor R, Attention (Vf, Val/Dis), Memory (S, V) |
| Right Motor | Age, Gender, Education, TIV, Motor L, Attention (Vf, Ave), Memory (S, V) | Gender, Lesion size, TIV, Language, Motor L, Attention (Vf, Ave, Val/Dis), Memory V | Age, Gender, Education, Lesion size, TIV, Language, Motor L, Attention (Vf, Ave, Val/Dis), Memory V |
| Spatial Memory | Age, Gender, Motor R, Attention (Vf, Val/Dis) | Age, Gender, Lesion size, Motor L, Attention (Vf, Val/Dis) | Gender, Lesion size, TIV, Motor L, Attention (Vf, Ave, Val/Dis) |
| Verbal Memory | Age, Gender, Lesion size, TIV, Inpatient (Ot, Pt, Slp), Motor (L, R), Attention (Vf, Ave, Val/Dis | Gender, Lesion size, TIV, Inpatient (Ot, Pt, Slp), Motor (L, R), Attention (Vf, Val/Dis) | Gender, Lesion size, TIV, Inpatient (Ot, Pt, Slp), Motor (L, R), Attention (Vf, Ave, Val/Dis) |
| Visual Fields  Attention | Age, Gender, Education, Lesion size, TIV, Language, Motor R, Attention (Ave, Val/Dis), Memory (S, V) | Age, Gender, Education, TIV, Inpatient (Ot, Pt, Slp), Language, Attention (Vf, Val/Dis), Memory (S, V) | Age, Gender, Education, Lesion size, TIV, Inpatient (Ot, Pt, Slp), Language, Motor (L, R), Memory (S, V) |
| Average Performance Attention | Age, Gender, Education, TIV, Inpatient (Ot, Pt, Slp), Motor R, Attention Vf, Memory V | Age, Gender, Education, Lesion size, Inpatient (Ot, Pt, Slp), Language, Motor (L, R), Attention (Vf, Val/Dis), Memory V | Gender, Education, TIV, Language, Motor R, Memory (S, V) |
| Validity / Disengagement Attention | Age, Gender, Education, Lesion size, TIV, Inpatient (Ot, Pt, Slp), Language, Motor L, Attention Vf, Memory (S, V) | Education, Lesion size, Inpatient (Ot, Pt, Slp), Language, Motor (L, R), Attention (Vf, Ave), Memory (S, V) | Age, Gender, Education, Lesion size, TIV, Inpatient (Ot, Pt, Slp), Language, Motor (L, R), Memory (S, V) |

Only covariates that were not significantly associated with the variables of interest were included in the analysis. Abbreviations: TIV, Total Intracranial Volume; OT, Occupational Therapy; PT, Physical Therapy; SLP, Speech-Language Pathology.

Supplementary Table 2**.** Linear mixed effects of deficits and recovery after stroke

|  | P-value（PCA-Scores） | | |
| --- | --- | --- | --- |
|  | Group | Post-stroke  time | Group&Time |
| Language | <0.001 | <0.001 | 0.0037 |
| Left Motor | <0.001 | <0.001 | 0 .8148 |
| Right Motor | <0.001 | <0.001 | 0.2577 |
| Spatial Memory | <0.001 | <0.001 | 0.0031 |
| Verbal Memory | <0.001 | <0.001 | 0.1603 |
| Visual Fields  Attention | 0.1197 | 0.2130 | 0.7084 |
| Average Performance Attention | 0.0090 | 0.0260 | <0.001 |
| Validity/Disengagement Attention | 0.0334 | 0.0608 | 0.4691 |

Supplementary Table 3 SSMN-deficit prediction models utilizing the Brainnetome Atlas (n = 246)

| Domain | two weeks | | three months | | one year | |
| --- | --- | --- | --- | --- | --- | --- |
|  | R  (Permutation  P) | MAE | R  (Permutation  P) | MAE | R  (Permutation  P) | MAE |
| Language | **0.54**  **(<0.001)** | 0.577 | 0.171  (0.095) | 0.607 | **0.256**  **(0.038)** | 0.671 |
| Left Motor | **0.59**  **(<0.001)** | 0.601 | **0.299**  **(0.02)** | 0.698 | **0.557**  **(<0.001)** | 0.588 |
| Right Motor | **0.476**  **(<0.001)** | 0.617 | **0.223**  **(0.05)** | 0.622 | **0.263**  **(0.033)** | 0.552 |
| Spatial Memory | **0.316**  **(0.012)** | 0.797 | 0.176  (0.108) | 0.809 | **0.328**  **(0.016)** | 0.766 |
| Verbal Memory | **0.269**  **(0.023)** | 0.813 | 0.113  (0.213) | 0.848 | 0.009  (0.326) | 0.87 |
| Visual Fields  Attention | 0.105  (0.206) | 0.57 | 0.193  (0.073) | 0.662 | 0.198  (0.009) | 0.5 |
| Average Performance Attention | 0.097  (0.214) | 0.72 | 0.067  (0.336) | 0.725 | - | - |
| Validity / Disengagement Attention | 0.024  (0.368) | 0.721 | - | - | 0.126  (0.207) | 0.702 |

Supplementary Table 4 SSMN-deficit prediction models demonstrated comparable performance to models employing other neuroimaging techniques

| Article | domain | model | Modality and metrics | Sample size | Model performance (*r^2^*) |
| --- | --- | --- | --- | --- | --- |
| Salvalaggio et al., 2020, Brain | (left and right visual, left and right motor, language, spatial attention, spatial and verbal memory | ridge regression | Voxelwise Lesion map | 25–116 | 0.06–0.58 |
|  |  |  | Lesion-based white matter disconnection maps | 25–116 | 0.05–0.42 |
|  |  |  | Rs-fMRI-based Functional disconnection maps | 25–116 | 0.012–0.38 |
| Siegel et al., 2016, PNAS | attention, visual  memory, verbal memory, language, motor, and visual | ridge regression | Voxelwise Lesion map | 100 | 0.11–0.65 |
|  |  |  | Rs-fMRI-based Functional connectivity | 100 | 0.13–0.51 |
| This study | attention, visual  memory, verbal memory, language, motor | Relevance vector regression | AAL-based SSMN | 83–103 | 0.05–0.41 |
|  |  |  | Brainnectome-based SSMN | 83–103 | 0–0.35 |

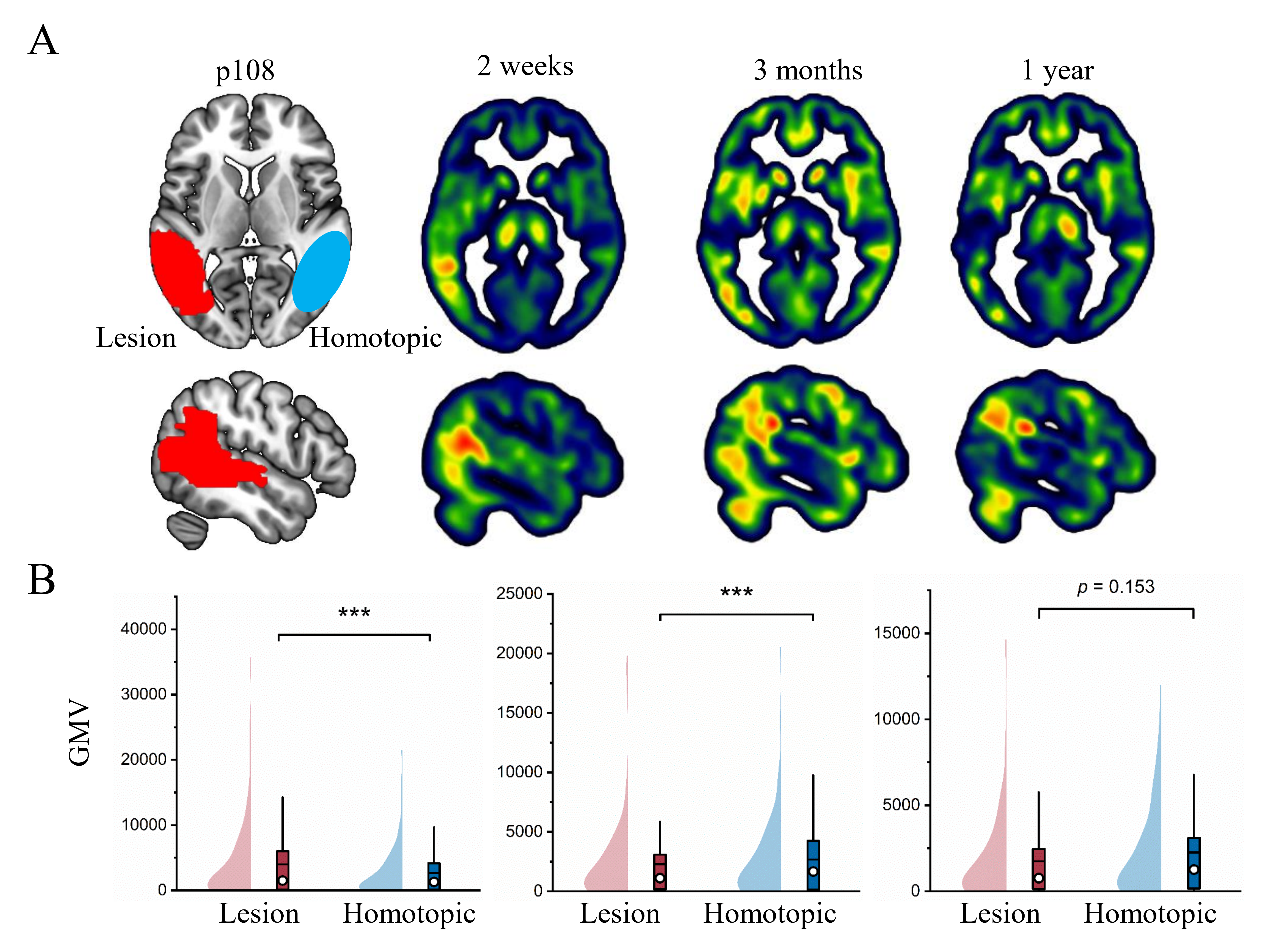

**Supplementary Figure 1**. **Normalization of GMV within stroke**. To construct the SSMN, we calculated the GMV without controlling for the stroke lesion. At 2 weeks and 3 months, the GMVs within the stroke lesion were significantly higher than those in homotopic areas. A: Whole-brain GMV for a single patient (patient 108) who suffered a large ischaemic stroke in the left posterior temporal and parietal areas. B. Paired t-test results between GMVs within the stroke lesion and those in homotopic areas. ***, *p* < 0.001.

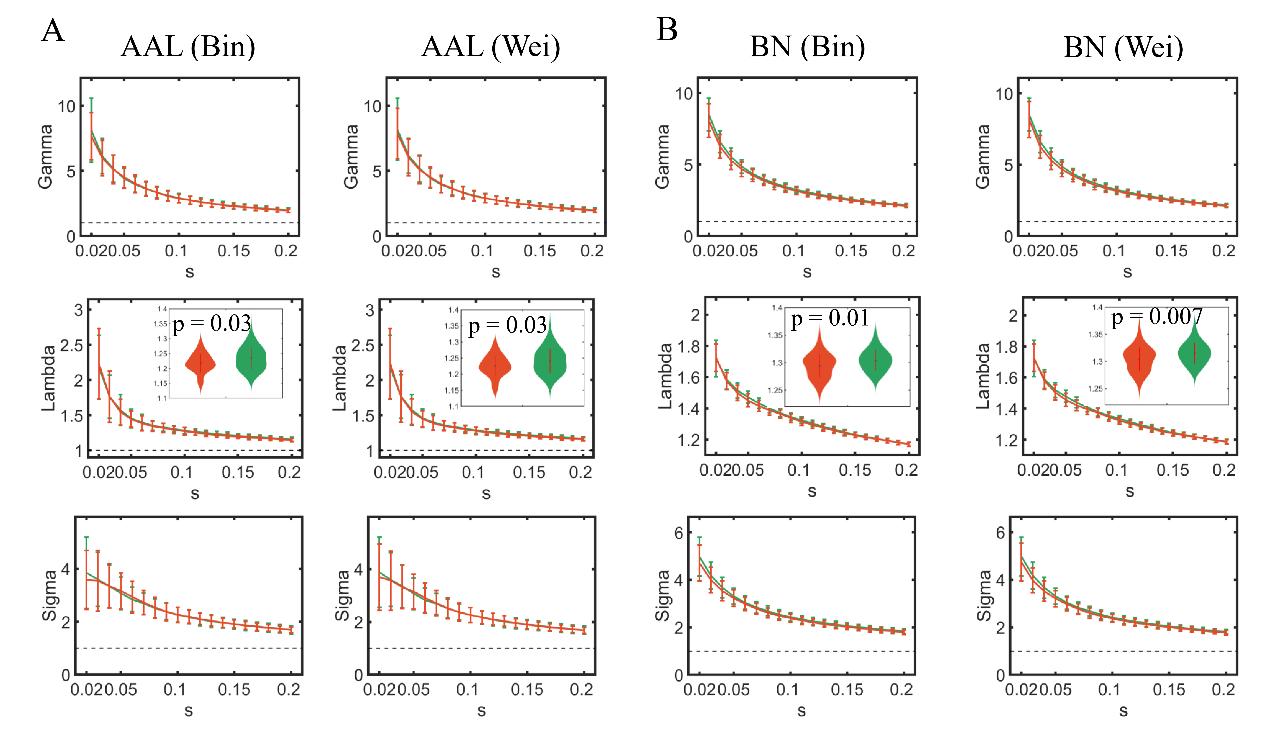

**Supplementary Figure 2**. **Disconnectome at two weeks after stroke assessed by SSMN for 103 patients (green)**. Significant increases in Lambda (*P* < 0.05, FDR correction) were observed for both AAL- and BN-based SSMN, when compared with HCs (red).

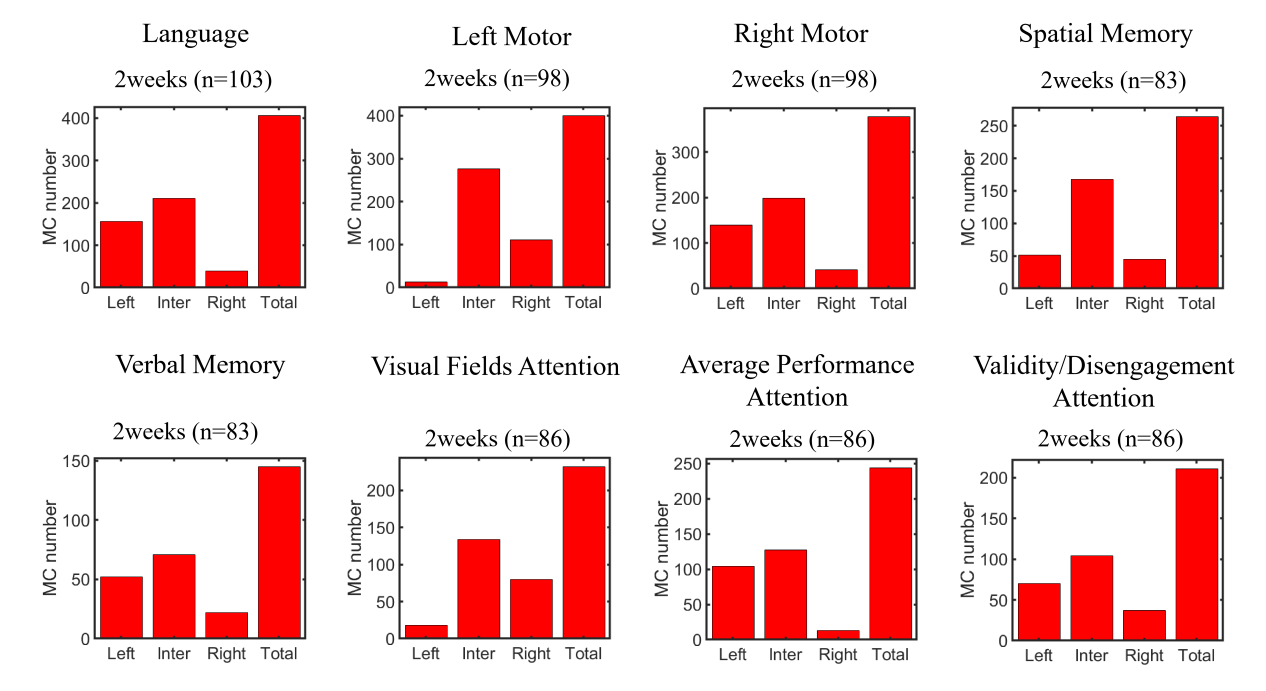

**Supplementary Figure 3**. **Types of morphological connections (MC) correlated with each domains at two weeks after stroke**. To assess the hemisphere asymmetry of correlation patterns, three sub-types of MCs were calculated: within left hemisphere (left), within right hemisphere (right), and inter-hemisphere (inter). For all domains, the inter-hemispheric MCs were dominant, with totally left-lateralized correlation patterns for domains of language, right motor, verbal memory, average performance attention and validity/disengagement attention, right-lateralized correlation patterns for domains of the left motor and visual fields attention, and symmetry correlation patterns for spatial memory.

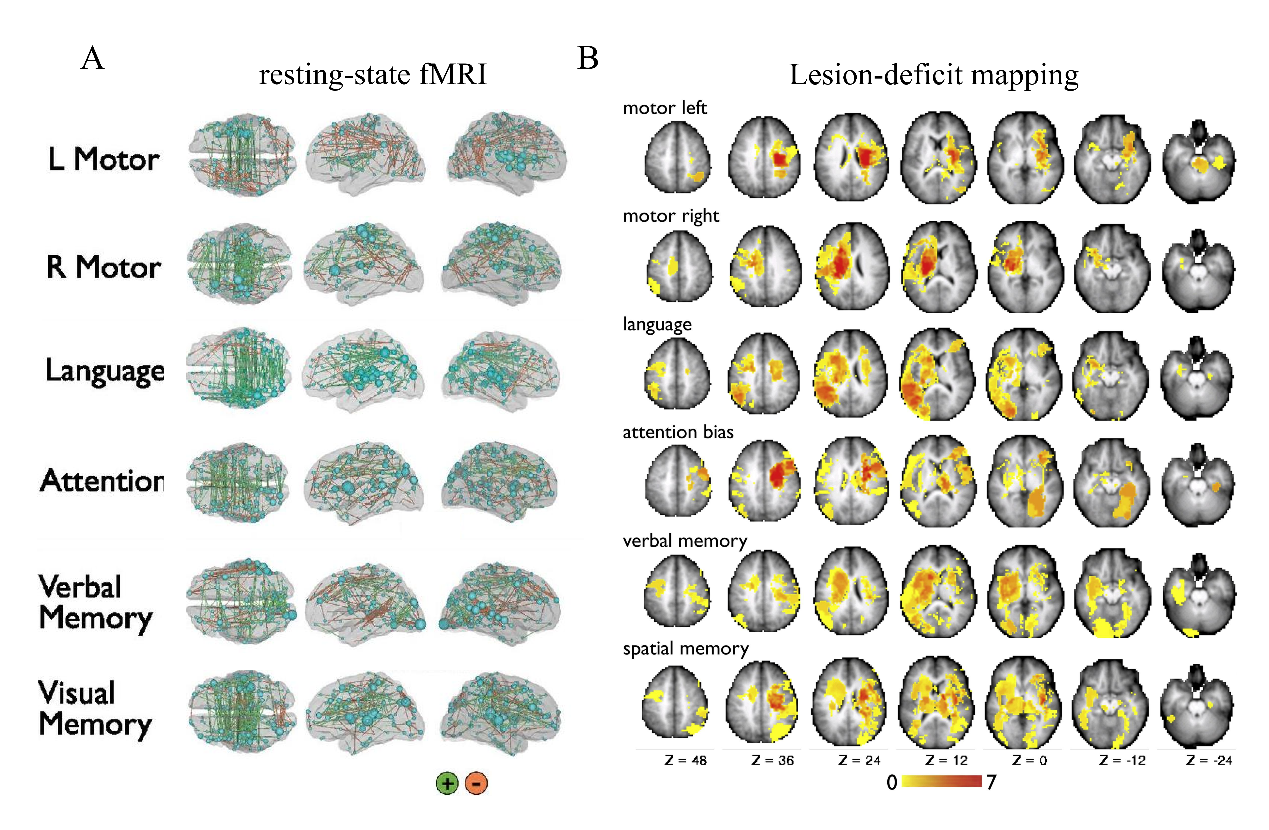

**Supplementary Figure 4**. **The rsFC-deficits correlation patterns in Siegel et al. (2016) and VLSM results in Corbetta et al. (2015)**. Note that the patient samples in these two studies were the same as those in this work. These results indicate that focal stroke leads to disconnection of white matter fiber bundles, disrupting both the brain's resting-state and individual morphological brain networks.

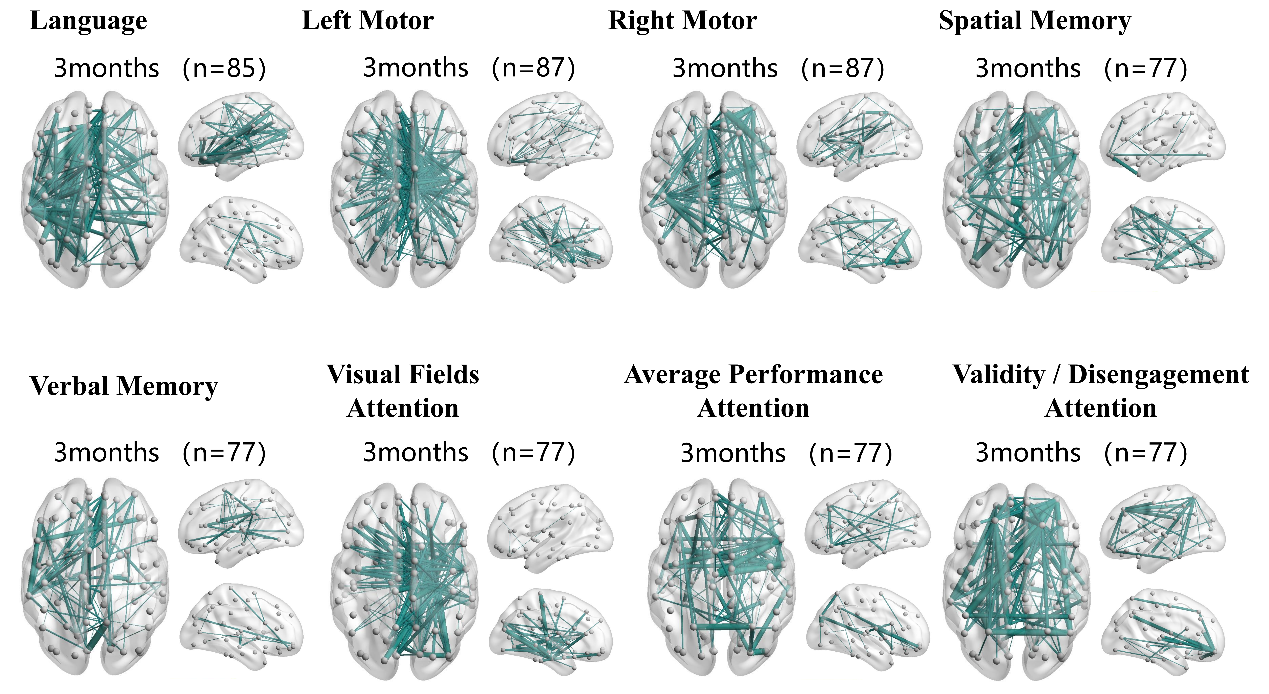

**Supplementary Figure 5. SSMN-deficits positive correlations at three months after stroke.** Edge *p*＜.05, component *p*＜.05 with NBS correction.

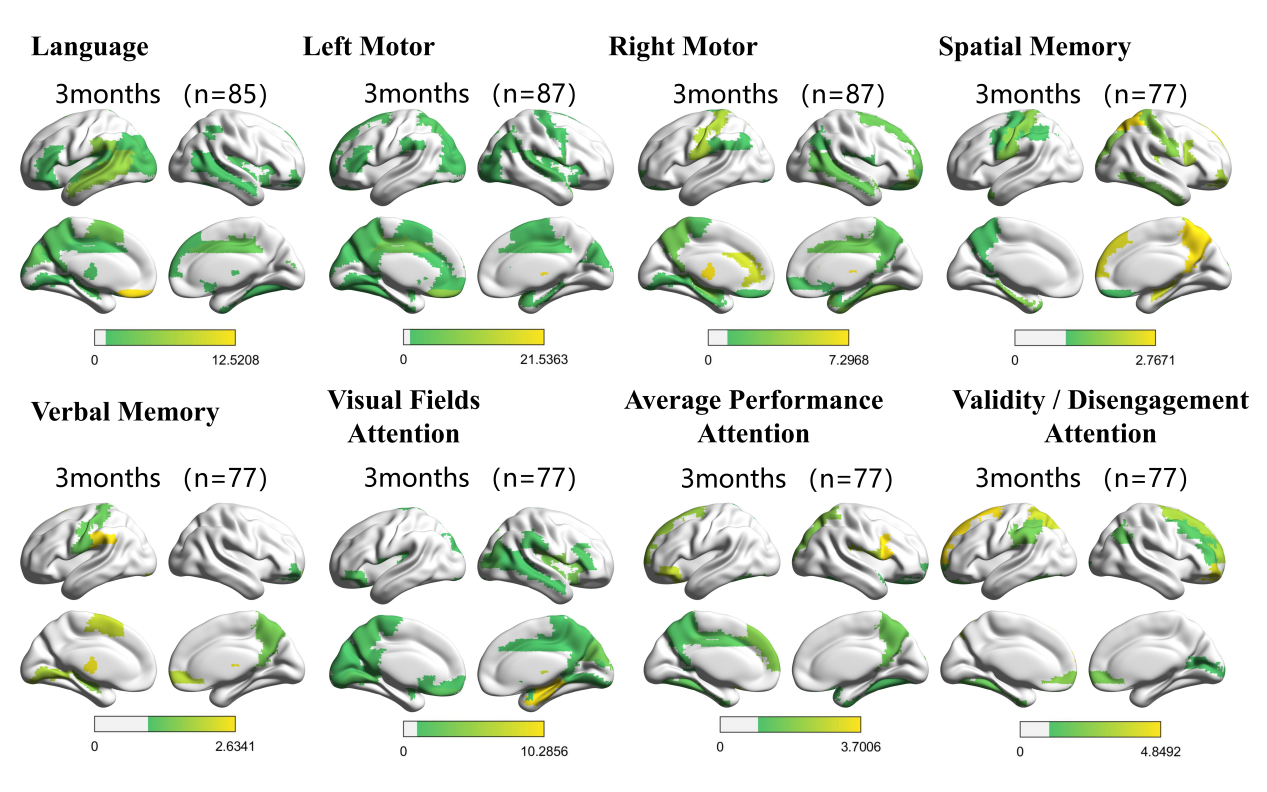

**Supplementary Figure 6**. Cortical renderings visualizing the SSMN connections associated with deficits at 3 months after stroke.

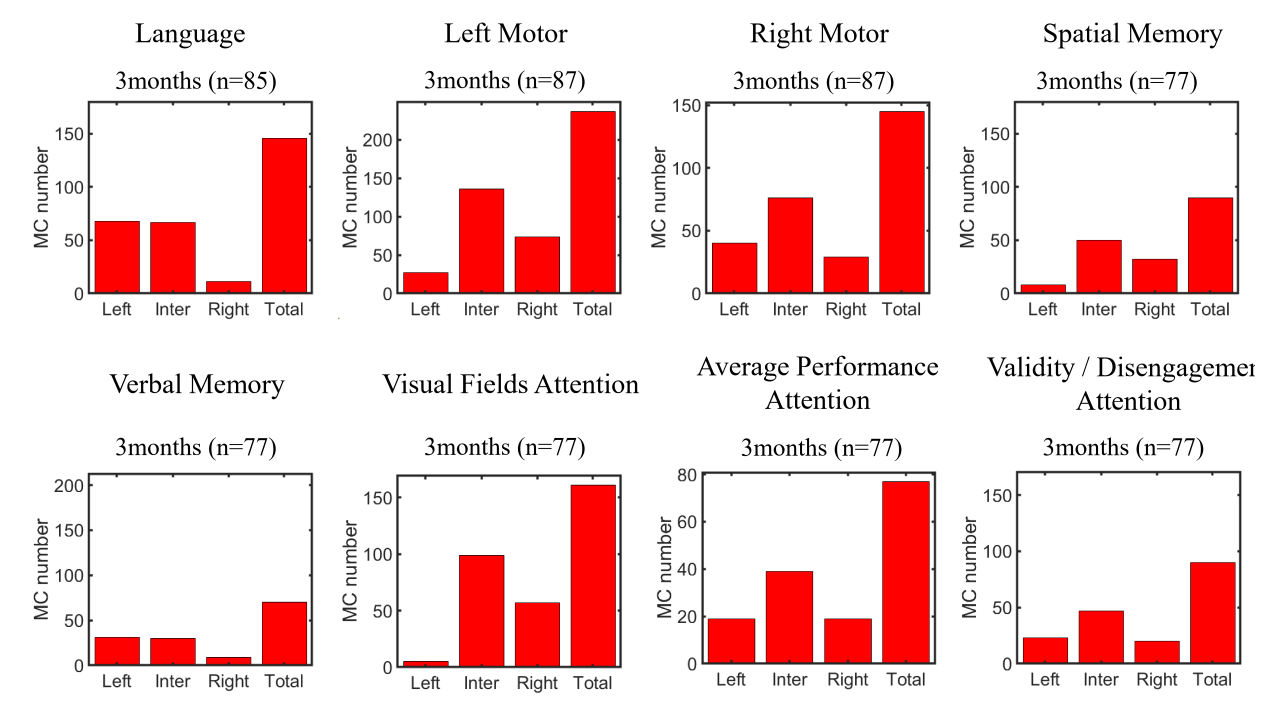

**Supplementary Figure 7**. **Types of morphological connections (MC) correlated with each domains at three months after stroke**. Left-lateralized correlation patterns were observed for domains of language, the right motor, verbal memory. Right-lateralized correlation patterns were observed for domains of the left motor, spatial memory and visual fields attention, and symmetry correlation patterns for average performance attention and validity/disengagement attention.

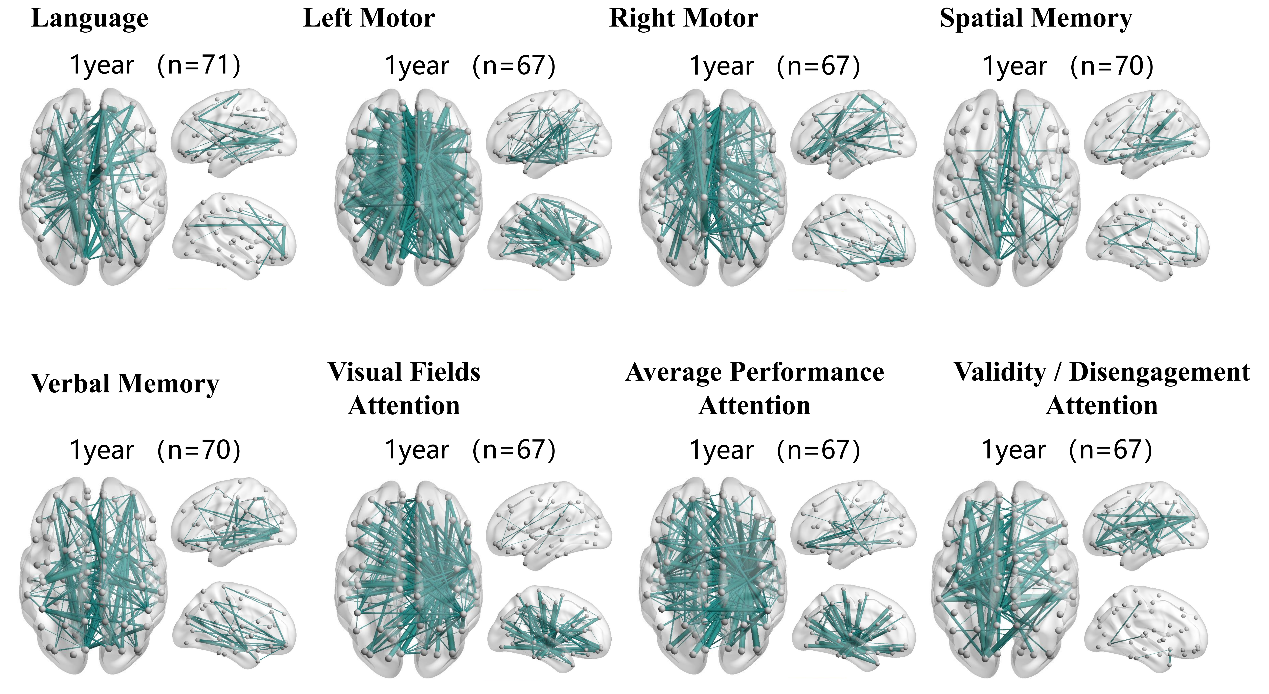

**Supplementary Figure 8. SSMN-deficits positive correlations at one year after stroke**. Edge *p*＜.05, component *p*＜.05 with NBS correction.

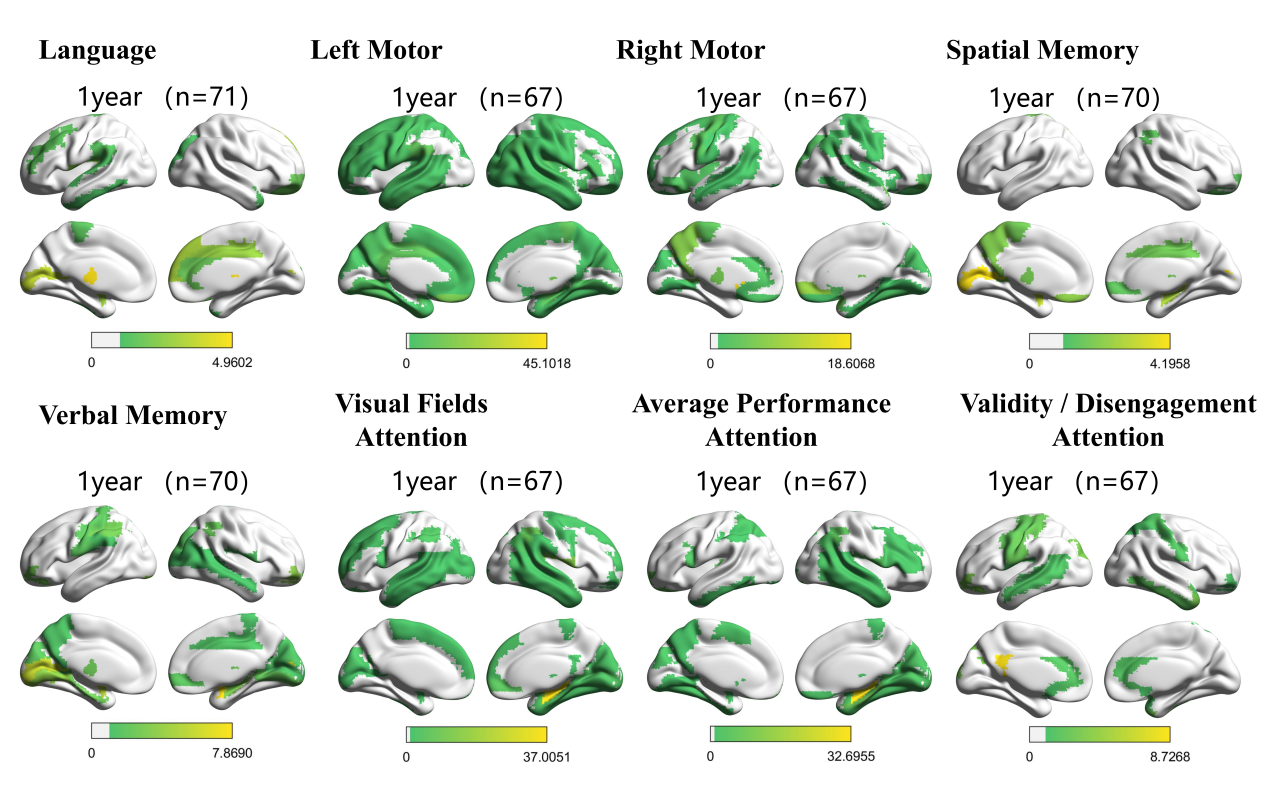

**Supplementary Figure 9**. **Cortical renderings visualizing the SSMN connections associated with deficits at one year after stroke**.

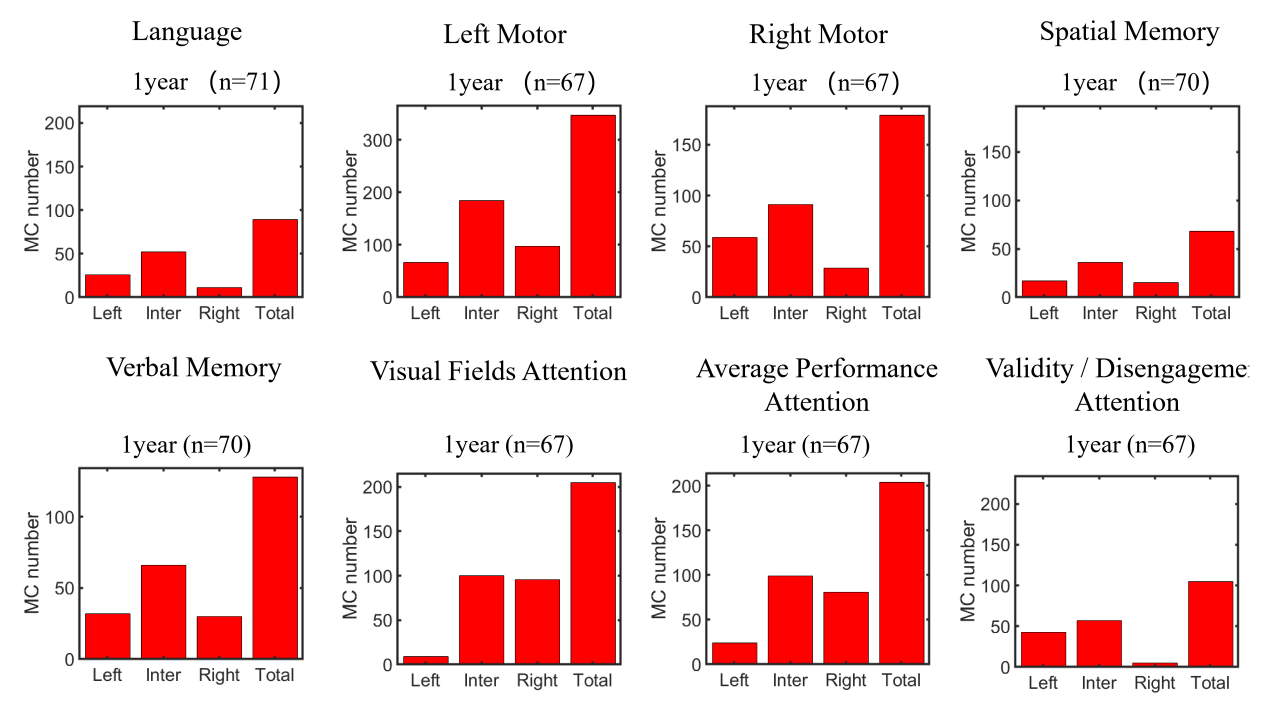

**Supplementary Figure 10**. **Types of morphological connections (MC) correlated with each domains at one year after stroke**. For all domains, the inter-hemispheric MCs were dominant, with totally left-lateralized correlation patterns for domains of language, right motor, visual fields attention, average performance attention, right-lateralized correlation patterns for domains of the left motor, validity/disengagement attention, and symmetry correlation patterns for domains of spatial and verbal memories.

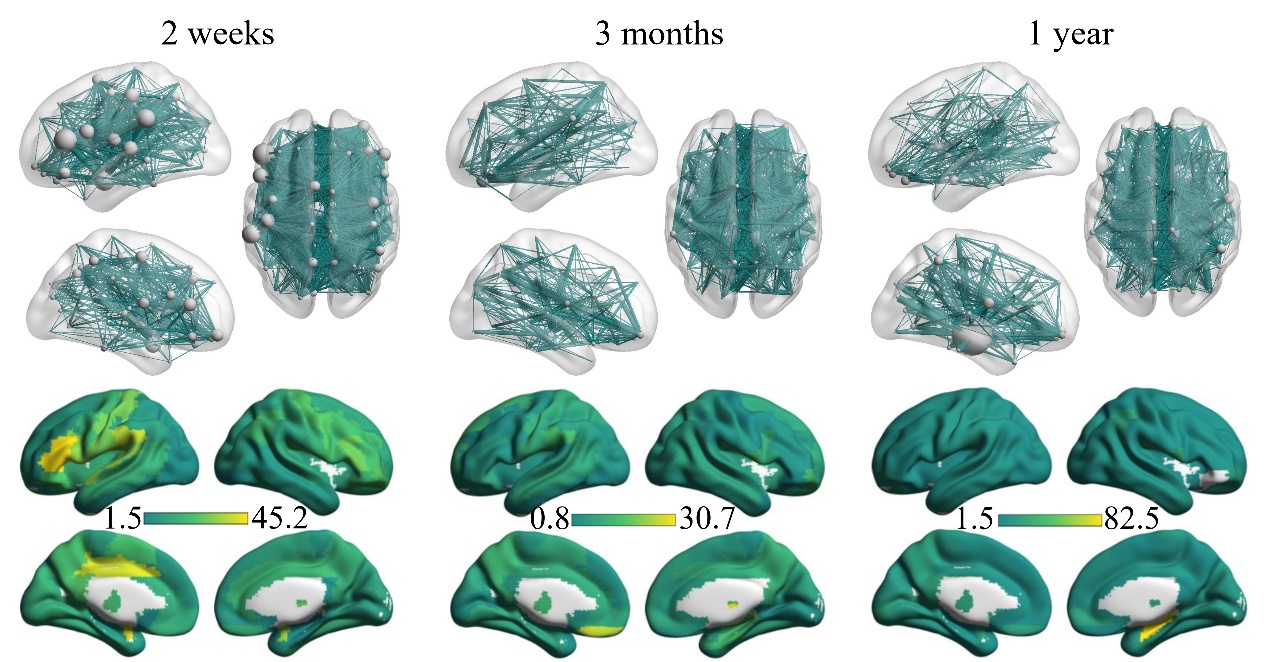

**Supplementary Figure 11**. **The overall SSMN-deficits correlations for all domains**. We summed the positive correlation values across all domains to assess which connections and brain regions contribute the most to post-stroke deficits. The size of the nodes in the figure is proportional to the total correlation value of the respective brain region.

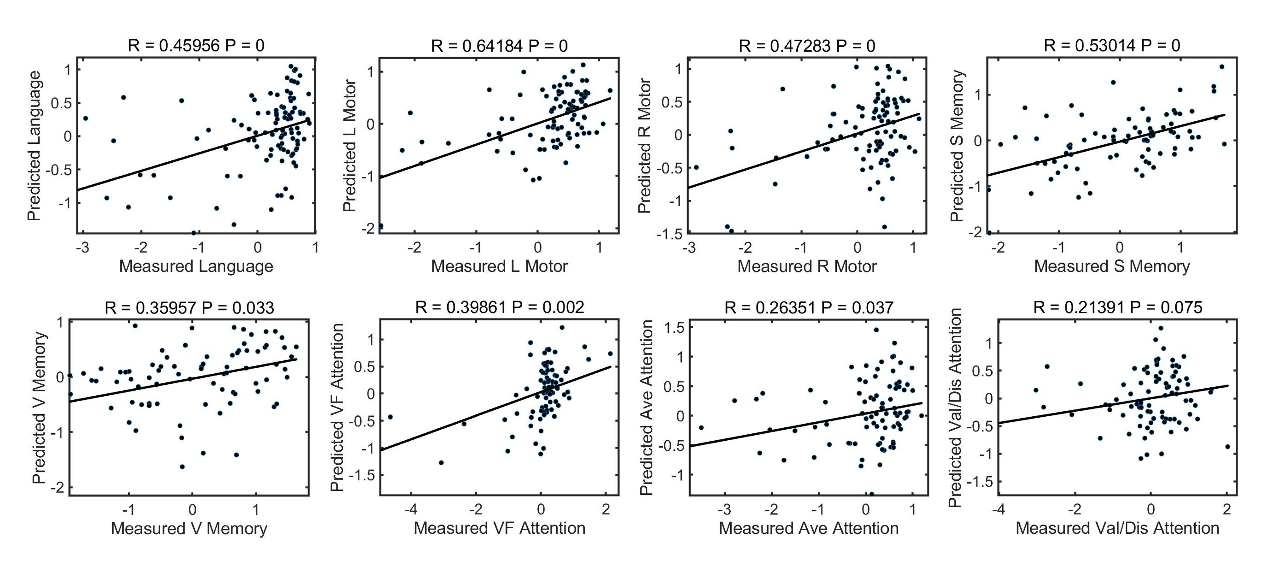

**Supplementary Figure 12**. **Scatter plots of AAL-based SSMN-deficits models two weeks post-stroke**. Patients’ SSMN significantly predicted individual patients’ behavioral/cognitive deficits across multiple domains. The solid lines represent the linear fitted lines between real and predicted scores. P = 0 means P < 0.001, i.e. no permutation correlation coefficient greater than the true prediction.

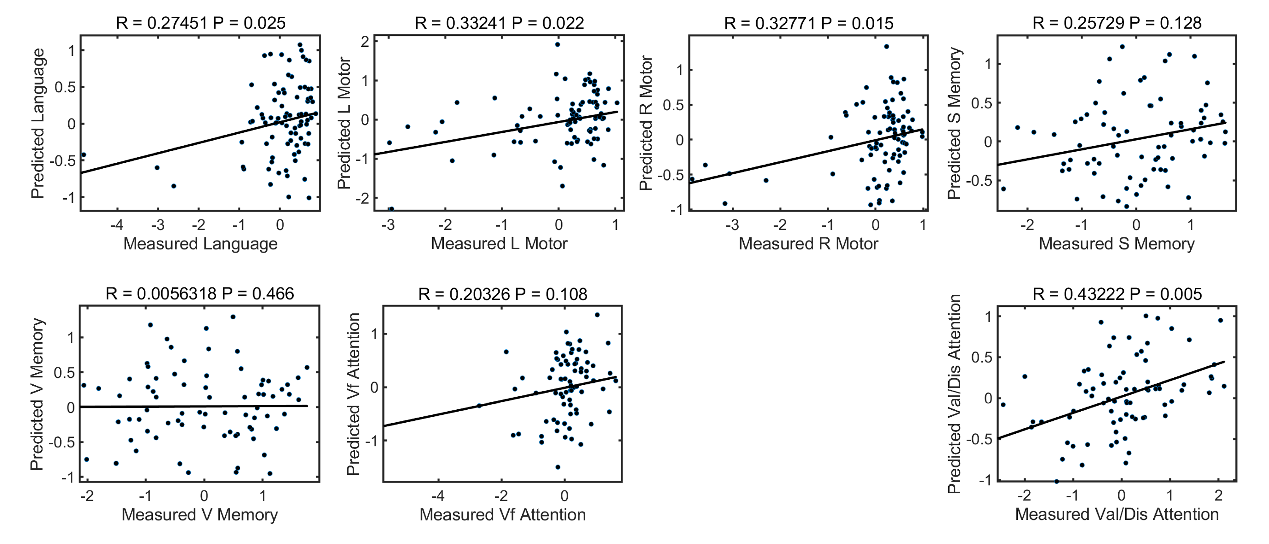

**Supplementary Figure 13**. **Scatter plots of AAL-based SSMN-deficits models 3 months post-stroke**. No predictive results were observed for average attention deficit.

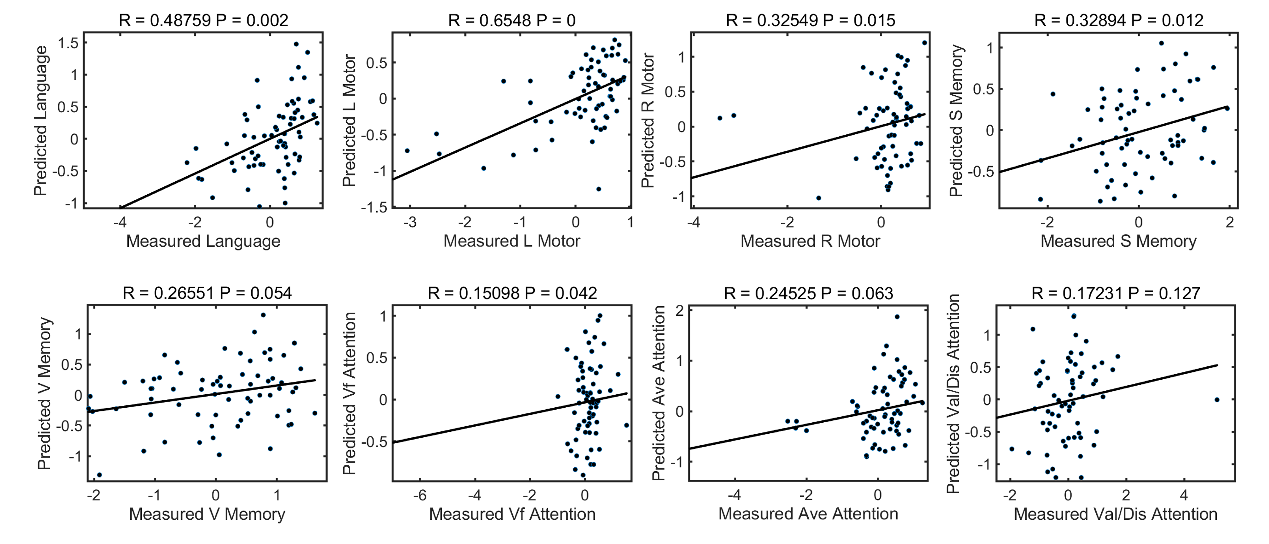

**Supplementary Figure 14**. **Scatter plots of AAL-based SSMN-deficits models one-year post-stroke**.

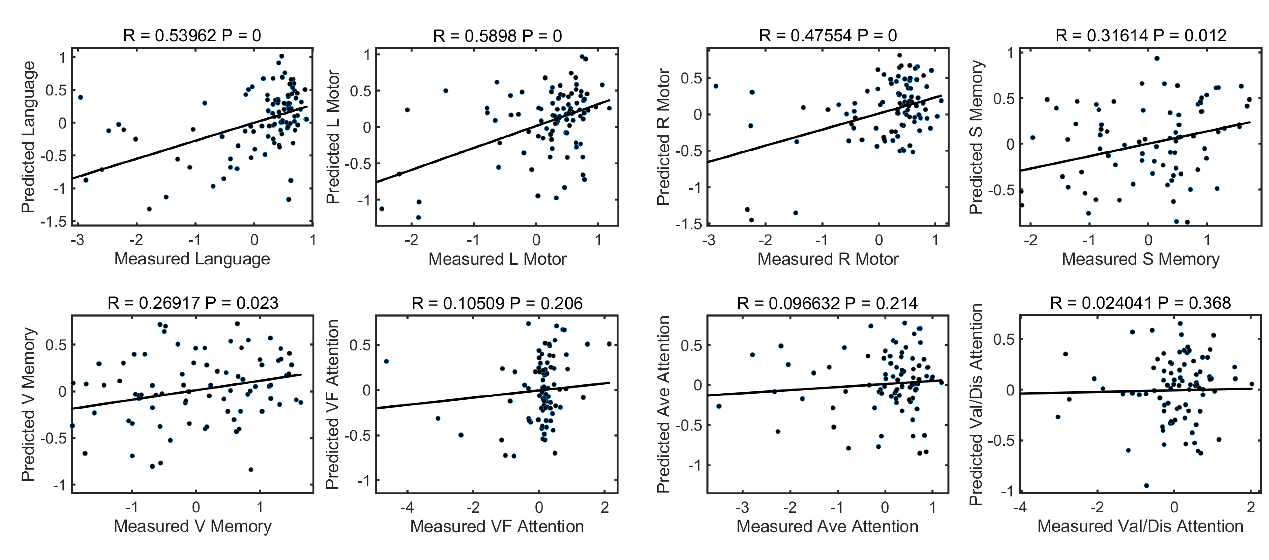

**Supplementary Figure 15**. Scatter plots of BN-based SSMN-deficits models two weeks post-stroke.

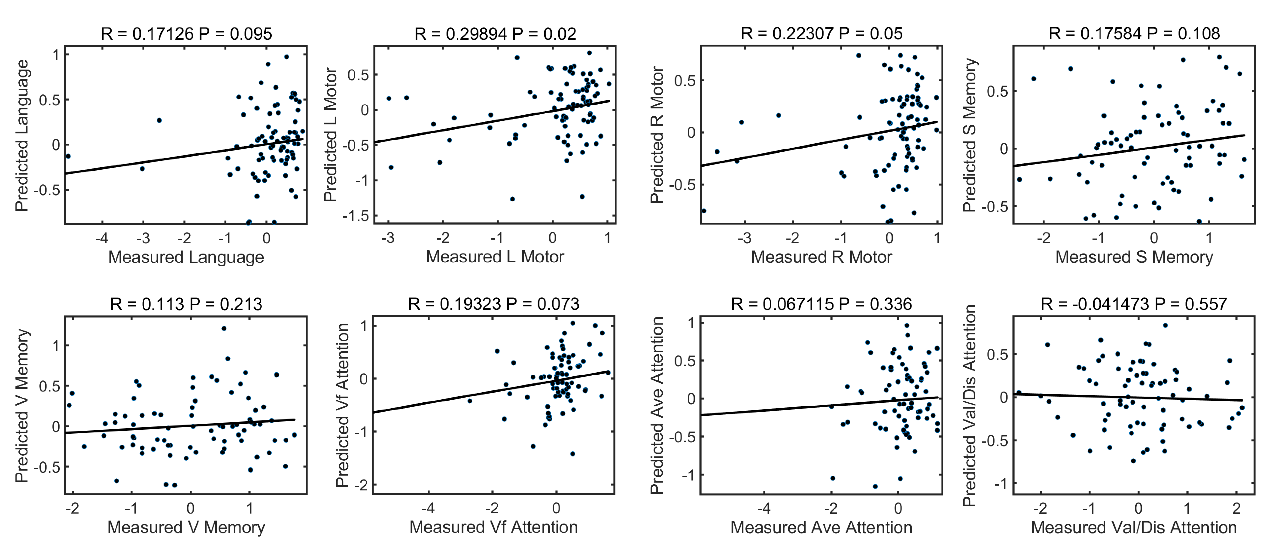

**Supplementary Figure 16**. Scatter plots of BN-based SSMN-deficits models three months post-stroke.

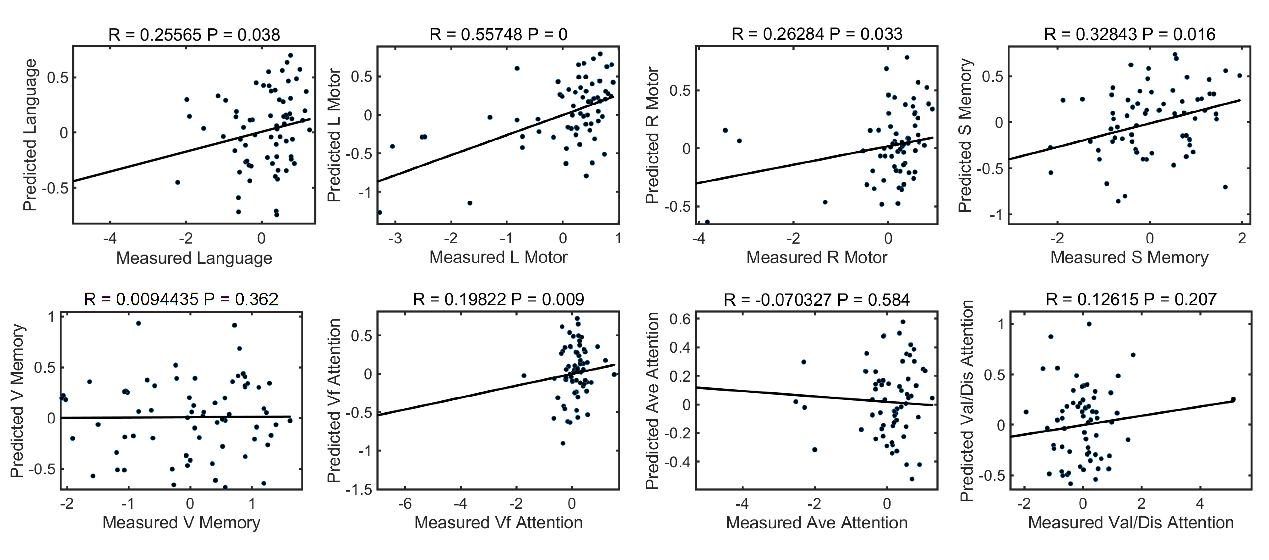

**Supplementary Figure 17**. Scatter plots of BN-based SSMN-deficits models one-year post-stroke.
